# From multiplicity of infection to force of infection in sparsely sampled high-transmission *Plasmodium falciparum* populations

**DOI:** 10.1101/2024.02.12.24302148

**Authors:** Qi Zhan, Kathryn E. Tiedje, Karen P. Day, Mercedes Pascual

**Author notes:** **For correspondence:** (QZ); (MP).

## Abstract

High multiplicity of infection (MOI), the number of genetically distinct parasite strains co-infecting a host, characterizes falciparum malaria and other infectious diseases under high transmission. High MOI in *Plasmodium falciparum* accompanies high prevalence of asymptomatic infection despite high exposure, creating a large transmission reservoir that challenges intervention. This pattern is enabled by parasite immune evasion through extensive antigenic diversity. The force of infection (FOI), the number of new infections acquired by an individual host over a given time interval, is the dynamic counterpart of MOI and a key epidemiological parameter for monitoring antimalarial interventions. FOI is difficult and costly to measure, especially in high-transmission regions, requiring cohort studies or model-based inference from repeated cross-sectional surveys. Here, we apply queuing theory to estimate FOI from MOI with two approaches: a two-moment approximation and Little’s Law. We illustrate these methods using MOI estimates obtained under sparse sampling schemes with the “*var*coding” approach. Both methods rely on infection duration data from naive malaria therapy patients and are therefore suitable for subpopulations with limited immunity, such as toddlers. We evaluate their performance using output from a stochastic agent-based model and apply the methods to an interrupted time-series study in northern Ghana, before and immediately after a three-round transient indoor residual spraying intervention. By accounting for sampling limitations with a Bayesian framework and bootstrap imputation, both methods yield good and replicable FOI estimates across various simulated scenarios. Their application to the surveys of 1-5-year-old children in Ghana indicates a larger than 70% reduction in annual FOI immediately after intervention.

## Introduction

Despite substantial intervention efforts, falciparum malaria in high-transmission regions remains a major public health concern, causing mortality among young children and a considerable economic burden, particularly in sub-Saharan Africa (***World Health Organization, 2023***). Thus, it remains important to robustly evaluate the effects of intervention efforts in these regions, including on transmission intensity. The force of infection (FOI), defined as the number of new *Plasmodium falciparum* infections acquired by an individual host over a given time interval, is a key metric reflecting the risk of infection and clinical episodes (***Mueller et al., 2012***). Whereas other metrics may describe the relationship between transmission intensity and the burden of malaria illness on global or continental scales (***Carneiro et al., 2010***; ***Beier et al., 1994***), FOI can relate local variation in malaria burden to transmission (***Mueller et al., 2012***). Although FOI is a key epidemiological parameter for malaria surveillance, it remains difficult, expensive, and labor-intensive to accurately measure, whether directly through cohort studies or indirectly through the fitting of epidemiological models. As molecular tools for parasite genomics become more readily available, they enable new approaches. In particular, molecular advances provide a basis for estimating a sister “static” quantity, the multiplicity of infection (MOI), defined as the number of genetically distinct parasite strains that co-infect a single human host (***Zhong et al., 2018***; ***Chang et al., 2017***; ***Ruybal-Pesántez et al., 2022***; ***Tiedje et al., 2022***; ***Labbé et al., 2023***). We can therefore ask whether we can go further, and on the basis of MOI obtain the dynamical, rate, quantity of force of infection.

Early efforts to directly measure FOI included clearing infections and observing the time to re-infection (***Macdonald, 1950***; ***Pull and Grab, 1974***; ***Msuya and Curtis, 1991***; ***Alonso et al., 2004***). Molecular approaches now enable the genotyping of individual parasite infections (***Mueller et al., 2012***; ***Hofmann et al., 2017***), but differentiating new infections from the temporary absence of an old infection in the peripheral blood and its subsequent re-emergence (***Daubersies et al., 1996***) remains challenging due to the low resolution of polymorphic markers and the complex within-host dynamics of malaria infection. Determining molecular FOI remains challenging, labor-intensive, and costly, requiring close long-term monitoring and genotyping of a large cohort.

Alternatives to direct measurements involve cross-sectional surveys with FOI estimated by fitting simple epidemiological models (***Bekessy et al., 1976***; ***Muench, 1959***; ***Smith and Vounatsou, 2003***; ***Felger et al., 2012***; ***Mugenyi et al., 2017***). These model-fitting procedures require empirical data sampled regularly and frequently, such as age-stratified large cohorts sampled six times a year, to account for FOI and infection duration heterogeneity (***Laishram et al., 2012***; ***Simpson et al., 2002***; ***Langhorne et al., 2008a***; ***Childs and Buckee, 2014***; ***Chang et al., 2016***; ***Ashley and White, 2014***) which is influenced by the interplay between host immunity and the antigenic composition of infections (***Piper et al., 1999***; ***Molineaux et al., 2002***; ***Doolan et al., 2009***; ***Barry et al., 2007***). Moreover, these approaches may face identifiability issues with model parameters (***Mugenyi et al., 2017***). Hence, these indirect measurements share limitations with direct ones.

Due to the described challenges, FOI has not become a readily available epidemiological quantity across geographical locations and times. In contrast, various approaches have been proposed to estimate MOI from clinical samples using size-polymorphic antigenic markers, microsatellites, and panels of biallelic single nucleotide polymorphisms (SNPs) (***Felger et al., 1994***; ***Konaté et al., 1999***; ***Anderson et al., 2000***; ***Daniels et al., 2008***; ***Chang et al., 2017***). Because *Plasmodium* parasites reproduce asexually during haploid stages within human hosts (***Guttery et al., 2012***), polymorphic genotypes indicate multiclonal infection. An alternative approach, termed *var*coding, leverages the extreme diversity of the *var* multigene family, which encodes the major variant surface antigen during blood-stage infection, and the resulting zero or extremely low similarity between repertoires with respect to their *var* gene composition shaped by immune selection. As methods for disentangling the molecular complexity of natural parasite infections across various transmission settings emerge and mature, MOI becomes more commonly and easily surveyed across space and time. Although MOI remains one of the most frequently used genetic metrics of parasite transmission (***Arnot, 1998***; ***Sondo et al., 2020***), it is by definition a number and not a rate.

A natural correlation exists between MOI and FOI, mediated by infection duration, which offers an opportunity to convert MOI into FOI. This conversion has been challenging because MOI estimates are often obtained from sparse sampling schemes, such as single-time-point surveys at the end of the wet (high-transmission) and dry (low-transmission) seasons (***Tiedje et al., 2022***; ***Abukari et al., 2019***). These MOI estimates have been useful (***Earland et al., 2019a***; ***Lee et al., 2006***) but the sparse sampling scheme behind them has limited their translation into transmission rates.

In this work, we propose using these MOI estimates for FOI inference with two mathematical modeling frameworks based on queuing theory (***Choi et al., 2005***; ***Little and Graves, 2008***). The two methods require infection duration values, for which we relied on data from naive malaria therapy patients with neurosyphilis (***Collins and Jeffery, 1999***; ***Maire et al., 2006***). Consequently, our approach is suited for FOI inference for subpopulations with a similar immune profile and the highest vulnerability, for example, infants or toddlers. We evaluate the methods through numerical simulation of an extended stochastic agent-based model (ABM) (***He et al., 2018***; ***Zhan et al., 2024***) for both closed and open systems with constant or seasonal transmission. We consider both homogeneous and heterogeneous transmission, with the latter including a high-risk group of hosts that receives the majority of the infectious bites. We also examine different statistical distributions for the times between local transmission events. We incorporate limitations representative of those encountered in the collection of field data into the sampling of simulation output, including under-sampling of *var* genes, missing data, and antimalarial drug treatment. We address these limitations in MOI estimates with a Bayesian framework and an imputation bootstrap approach. Both methods provide good and replicable FOI estimates across simulated scenarios. After validating with simulations, we apply the two methods to empirical data from an interrupted time-series study in Bongo District, northern Ghana, that involved a three-round transient indoor residual spraying (IRS) intervention (***Tiedje et al., 2022, 2025***). We focus on children aged 1-5 years whose immune profiles are closer to naive patients than the rest of the population, an aspect we discuss later. We then explore the relationship between FOI and another commonly used surrogate for transmission intensity, the entomological inoculation rate or EIR, defined as the number of infectious bites received by an individual over a given time period (***Shaukat et al., 2010***). This relationship underscores the challenges of relating measures of transmission intensity to malaria burden at local scales and achieving substantial reductions in transmission in high-transmission regions.

## Results

### The Bayesian formulation of the *var*coding method, combined with the bootstrap imputation approach, effectively addresses sampling limitations often encountered in collecting field data for MOI estimates

Because our FOI inference relies on MOI estimates, we first investigate the impact of various sampling limitations on these estimates. We use the Bayesian formulation of the *var*coding method and the bootstrap imputation approach (Materials and Methods) to address sampling limitations often encountered in collecting field data for MOI estimates: under-sampling or imperfect detection of *var* genes, missing data, antimalarial drug treatment, and their combination. For this investigation, we utilize simulation output from an agent-based model (ABM) of malaria transmission with known true MOI values (***He et al., 2018***; ***Zhan et al., 2024***). Key assumptions and processes for the ABM, and the experimental design for simulation output are summarized in Appendix 1-Simulation data, with illustrations in Appendix 1-Figure 5-6.

Our results indicate that MOI estimates obtained using the Bayesian formulation of the *var*coding method and the bootstrap imputation approach closely match true MOI values in most cases. To assess the difference between estimated and true MOI distributions, we use the Cramer-von Mises and Anderson-Darling tests. The Cramer-von Mises test quantifies the sum of the squared differences between cumulative distribution functions, while the Anderson-Darling test, a modification of the former, gives more weight to the tails of distributions. Most p-values are non-significant (> 0.05), indicating insufficient evidence to conclude that the estimated and true distributions differ, with few exceptions in pre-IRS or low-coverage IRS scenarios. The Bayesian formulation of *var*coding tends to underestimate MOI because it assumes that each co-infecting strain contributes a distinct set of *var* genes. In practice, limited overlap among co-infecting strains reduces the number of *var* genes detected per individual relative to this expectation, thereby leading to systematic underestimation of MOI. This underestimation bias can be more pronounced in certain high-transmission situations where many hosts have a high true MOI, such as the aforementioned exceptions in pre-IRS or low-coverage IRS scenarios. Consequently, this underestimation in MOI leads to an underestimation of FOI estimates, as described in the next section. Detailed results of both tests are provided in the supplementary file 1-MOImethodsPerformance.xlsx.

The distributions of MOI estimates across different surveys in Ghana and from the simulated outputs are not Poisson-distributed (Appendix 1-Test of deviation from Poisson homogeneity in MOI estimates, supplementary file 2-deviationFromPoissonTest.xlsx) (***Potthoff and Whittinghill, 1966***; ***Lloyd-Smith et al., 2005***). This deviation suggests that infection arrivals depart from a homogeneous Poisson process. In addition, infection durations often deviate from an exponential distribution, violating the assumptions under which Poisson MOI arises. Together, these departures complicate conversion from MOI to FOI in the presence of a finite host carrying capacity, but the proposed methods are flexible and applicable (Materials and Methods-Inferring FOI from MOI estimates).

### The Two-Moment Approximation and Little’s Law methods give good and replicable estimates for FOI across various simulated scenarios

We begin with a homogeneous exposure risk scenario for seasonal transmission in a closed system (Appendix 1-Figure 6A-C, and Appendix 1-Simulation data). The times between local transmission events follow a Gamma distribution (Appendix 1-Figure 6C). We infer FOI using the true MOI values and the MOI estimates obtained via the Bayesian formulation or the bootstrap imputation approach, with each method accounting for the specific sampling limitations applied in that scenario, whether a single limitation or all limitations combined. Details on deriving the confidence intervals are provided in Appendix 1-Confidence intervals for FOI inference.

Across pre-IRS and three IRS coverage levels, the 95% confidence intervals and the bootstrap distributions of FOI estimates are narrow. FOI estimates based on the true MOI values, as well as those based on MOI estimates obtained under (i) the missing data limitation and (ii) the antimalarial treatment limitation, each accounted for by the bootstrap imputation approach, closely match the true FOI values (Figure 1). FOI estimates based on MOI estimates obtained under (i) the under-sampling or imperfect detection of *var* genes and (ii) all sampling limitations combined, accounted for by the Bayesian formulation alone in the former case and by the Bayesian formulation together with the bootstrap imputation approach in the latter, show slight underestimations (Figure 1), due to the underestimation of MOI described in the previous section.

**Figure 1.**
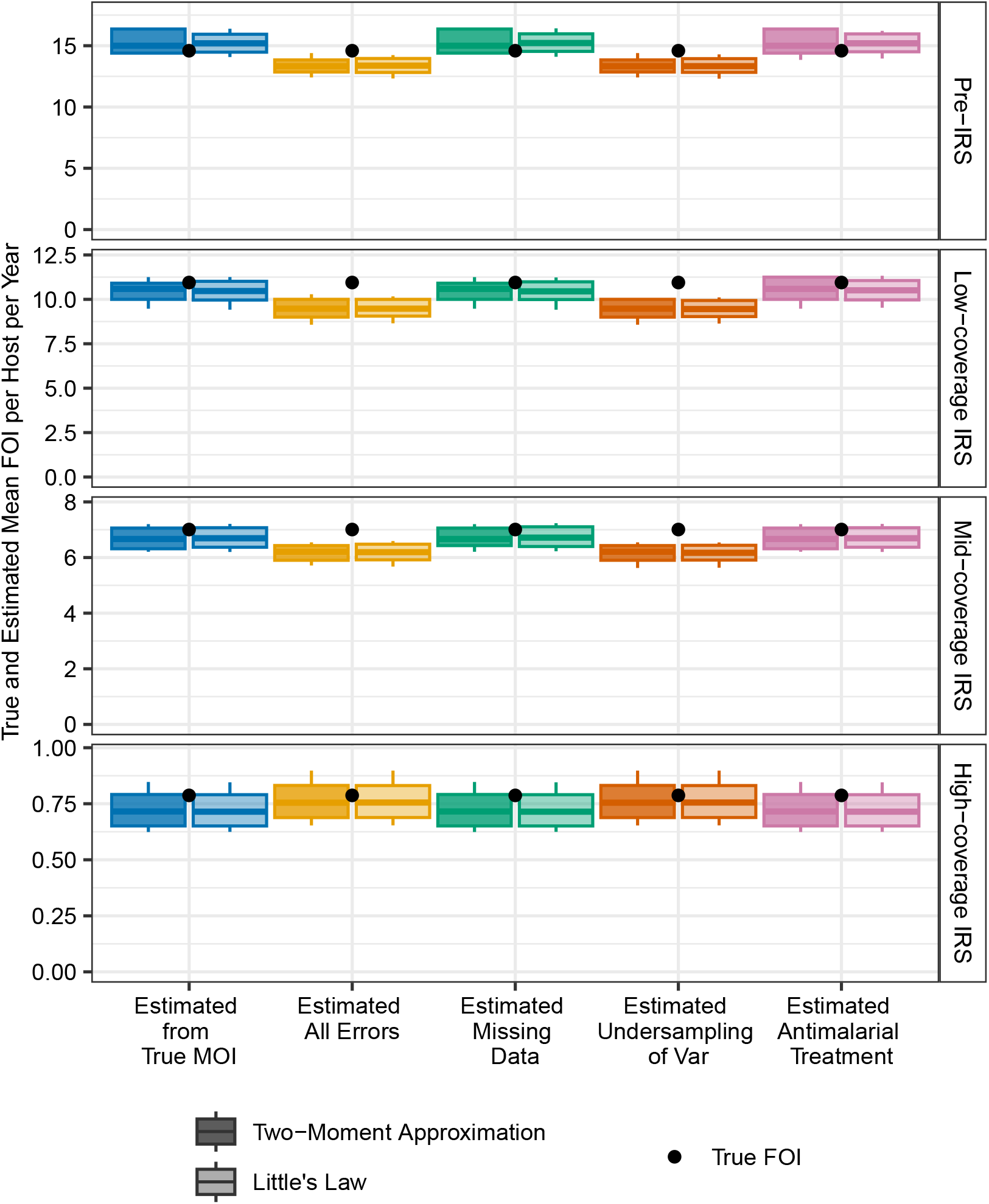
Confidence intervals for estimated mean FOI values in simulated scenarios of homogeneous exposure risk, before and during IRS interventions at three different coverage levels. The times between local transmission events follow a Gamma distribution, with seasonal transmission in a closed system. FOI estimates are derived from true MOI values and MOI estimates obtained through the Bayesian formulation or the bootstrap imputation approach correcting for all or individual sampling limitations. The true mean FOI per host per year is computed by dividing the total number of infections acquired by the population by the total number of hosts in the population. Each boxplot shows minimum, 5% quantile, median, 95% quantile, and maximum values.

To quantify the difference between inferred and true FOI values, we check if the true FOI lies within the bootstrap distribution and calculate the relative deviation, which is defined as the true FOI value minus the median of the bootstrap distribution for the estimate, normalized by the true FOI value. These details are in supplementary file 3-FOImethodsPerformance.xlsx.

We continue with a heterogeneous exposure risk scenario in which a high-risk group (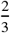 of the population) receives approximately 94% of bites, while a low-risk group (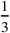 of the population) receives the remainder (Appendix 1-Figure 6C). Transmission is seasonal and the system is semi-open (Appendix 1-Figure 6A-B, and Appendix 1-Simulation data). The times between local transmission events are Gamma-distributed (Appendix 1-Figure 6C). As before, FOI estimates across pre-IRS and three IRS coverage levels show narrow 95% confidence intervals and bootstrap distributions close to true FOI values (Figure 2), with slight underestimations for FOI estimates based on MOI estimates corrected for under-sampling or imperfect detection of *var* genes and those corrected for all sampling limitations.

**Figure 2.**
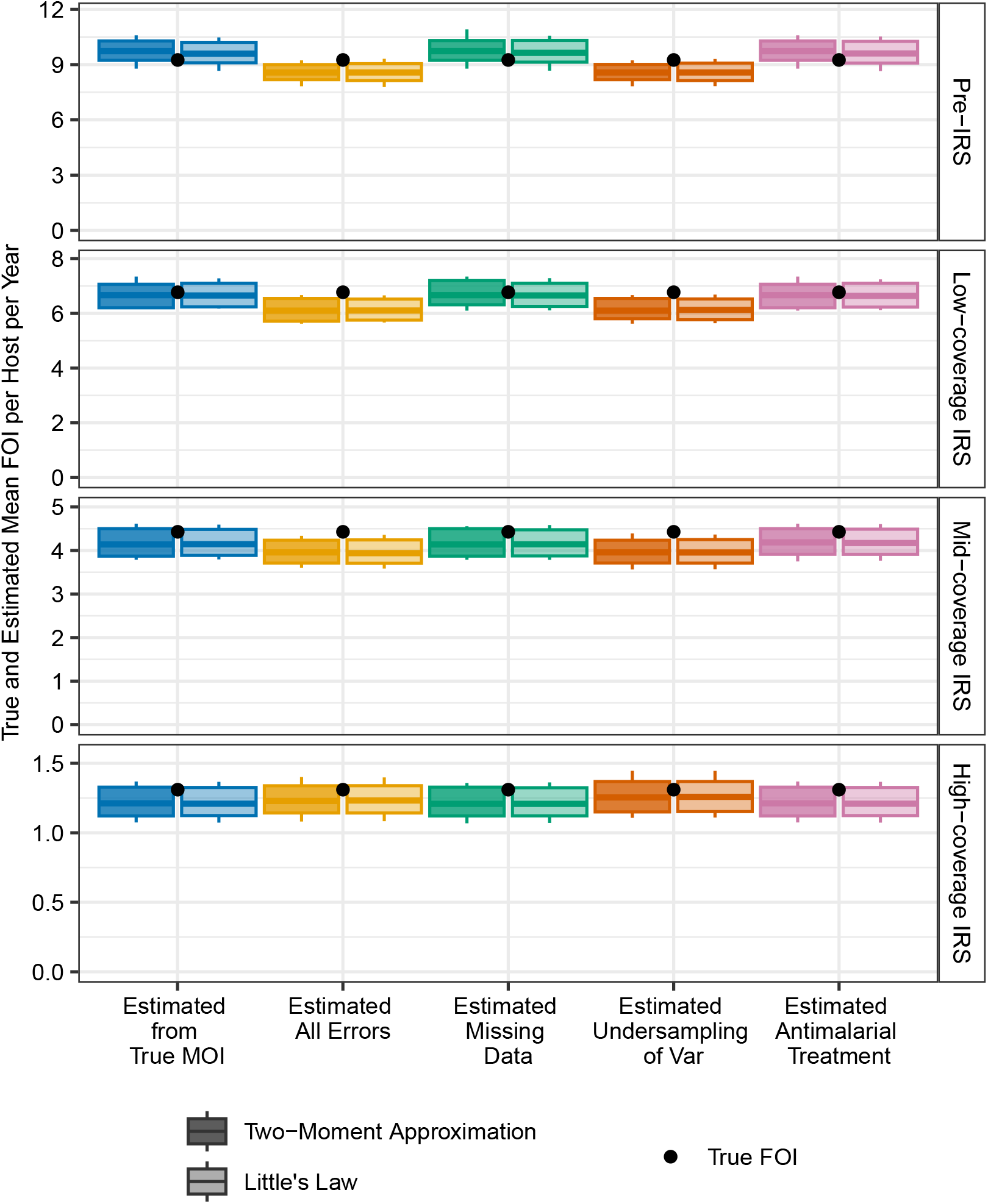
Confidence intervals for estimated mean FOI values in simulated scenarios of heterogeneous exposure risk, before and during IRS interventions at three different coverage levels. The times between local transmission events follow a Gamma distribution, with seasonal transmission in a semi-open system. FOI estimates are derived from true MOI values and MOI estimates obtained through the Bayesian formulation of the *var*coding method or the bootstrap imputation approach correcting for all or individual sampling limitations. The true mean FOI per host per year is computed by dividing the total number of infections acquired by the population by the total number of hosts in the population. Each boxplot shows minimum, 5% quantile, median, 95% quantile, and maximum values.

The performance of the two methods across additional simulated scenarios is shown in Appendix 1-Figure 11-22.

### The Two-Moment Approximation and Little’s Law methods give replicable FOI estimates for empirical surveys conducted in the Bongo District of northern Ghana

After validating with simulations, we apply the two methods to empirical surveys in the Bongo District of northern Ghana. We first derive their MOI estimates. Due to the high but imperfect detection power of PCR, we assume three levels of sensitivity: 0% (high detectability), 5% (mid detectability), and 10% (low detectability) of PCR-negative individuals carrying infection (Materials and Methods-The under-sampling of infections in empirical surveys).

Antimalarial treatment, sought in response to symptoms or perceived transmission risk, can impact the duration of an ongoing infection and may therefore violate the assumption underlying the two methods, which rely on infection duration data from naive malaria therapy patients with neurosyphilis. We address this issue by either excluding treated individuals from the analysis or by discarding their infection status and MOI estimates, instead sampling from non-treated individuals with MOI > 0. Since the latter samples non-zero MOIs for these treated and uninfected individuals, it results in an upper bound for FOI estimates. Note that in this latter case, we do not assume that the MOI distribution for treated individuals is the same as that for untreated individuals. Rather, we aim to estimate what their MOI would have been, and consequently, determine what the FOI per individual per year in the combined population would be, had these individuals not received antimalarial treatment. Further details can be found in Materials and Methods-Antimalarial drug treatment of infections in empirical surveys.

Next, we apply the two-moment approximation and Little’s Law methods to derive FOI estimates from empirical MOI estimates. Both methods yield replicable FOI estimates. The FOI estimates are similar across the three PCR sensitivity levels and the two approaches for handling treated individuals. The 95% confidence intervals and the full sampling distributions from bootstrap analysis are concentrated (Figure 3, Appendix 1-Figure 23-25). Notably, there is a significant reduction in FOI, exceeding 70%, indicating that the three-round IRS intervention, although transient, was highly effective.

**Figure 3.**
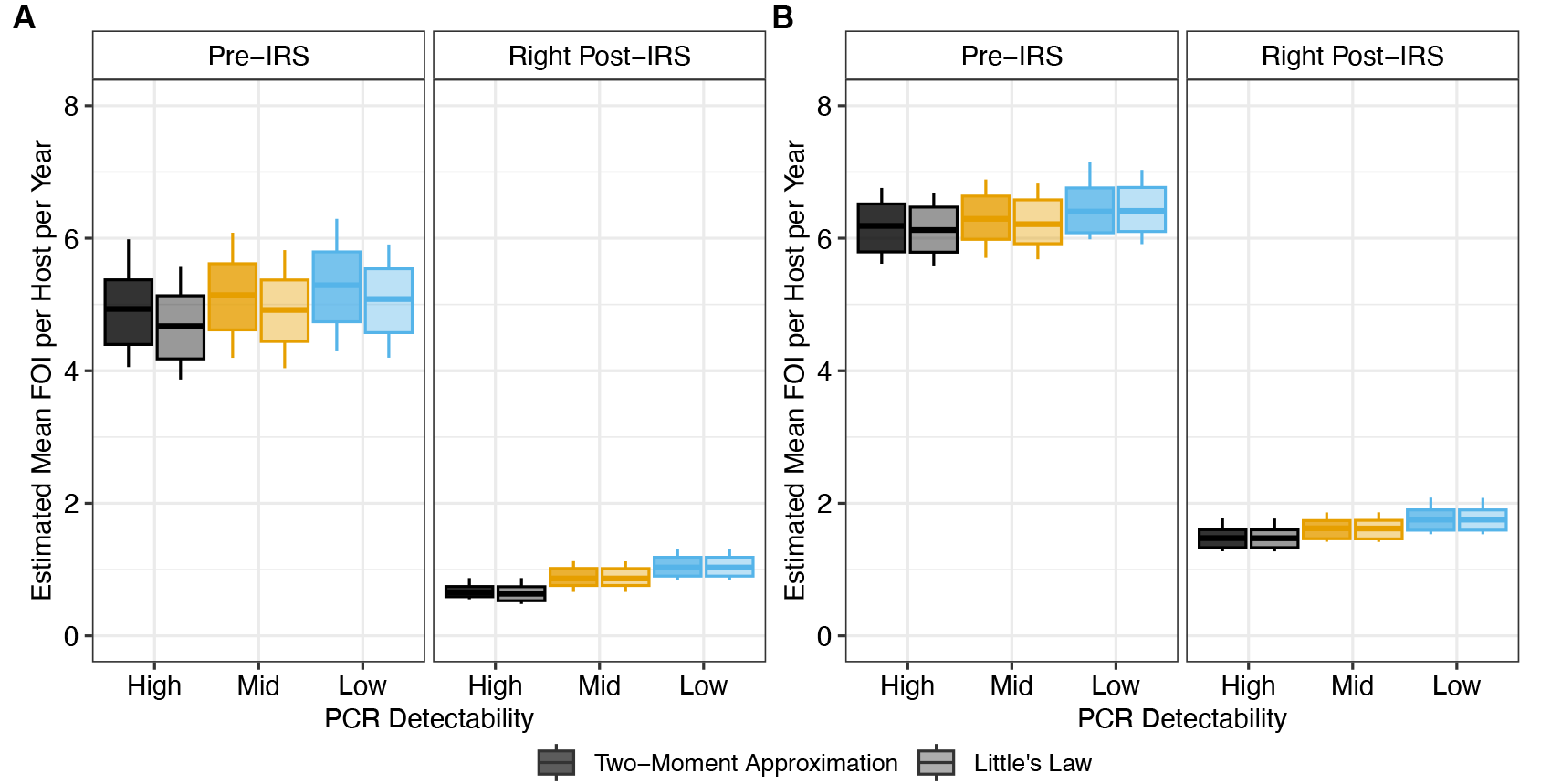
Confidence intervals for the estimated mean FOI values in Ghana surveys before and immediately after a transient three-round IRS intervention. (A) The estimated FOI values when excluding these treated individuals from the analysis. (B) The estimated FOI values when discarding the infection status and MOI estimates of treated individuals and sampling from non-treated ones with MOI > 0. Since this case samples non-zero MOIs for these treated and uninfected individuals, it results in an upper bound for FOI estimates. Each boxplot shows minimum, 5% quantile, median, 95% quantile, and maximum. The value of *c* is set to 30. FOI estimates with other values of *c* can be found in Appendix 1-Figure 23-25.

### The inferred FOI and directly measured EIR from the Ghana surveys align with the relationship between these two quantities in previous studies

We plot the measured annual EIR against the estimated annual FOI, and the transmission efficiency (the ratio between FOI and EIR) against the measured annual EIR from previous field studies summarized by (***Smith et al., 2010***) (Figure 4). In these studies, FOI estimation was based on fitting a simple epidemiological model to age-stratified prevalence data from cross-sectional parasitological studies (***Smith et al., 2010***; ***Pull and Grab, 1974***), while EIR was estimated using various methods such as exit bait collection, human bait collection, pyrethrum spray collection, night bite collection, and outdoor resting collection (***Smith et al., 2010***). The yellow line indicates the functional curve fitted to these data points (Materials and Methods-Conversion between FOI and the Entomological Inoculation Rate, or EIR), initially proposed by (***Smith et al., 2010***).

**Figure 4.**
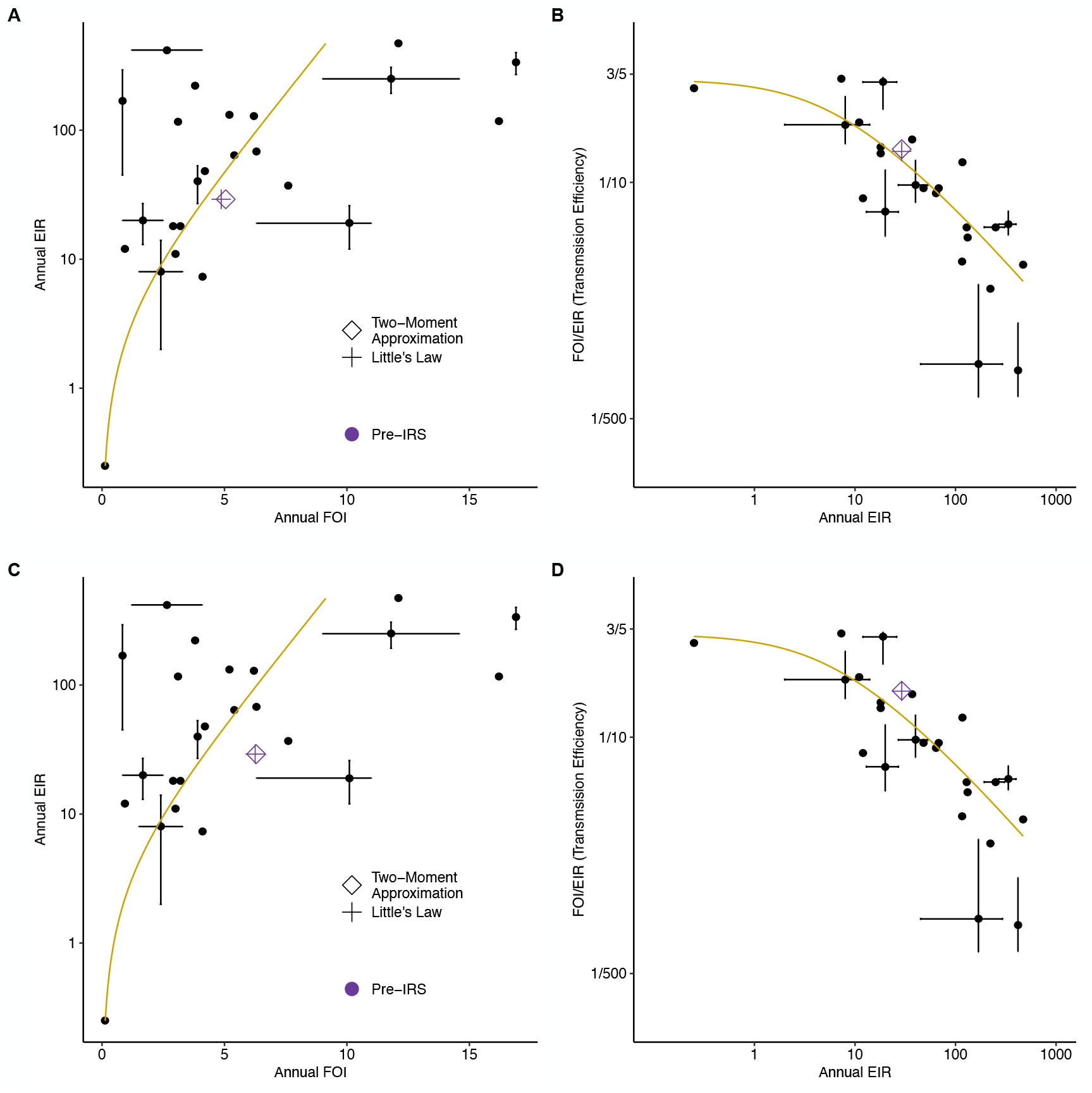
The saturation in FOI with increasing EIR and their non-linear relationship from previous field studies. (A) and (B) present our empirical estimates (with *c* = 30) when excluding treated individuals from the analysis. (C) and (D) show our estimates when discarding the infection status and MOI estimates of treated individuals and instead sampling from non-treated ones with MOI > 0. Since this case samples non-zero MOIs for these treated and uninfected individuals, it results in an upper bound for FOI estimates. The black points represent paired EIR-FOI values from the literature, as summarized by (***Smith et al., 2010***), with crosses indicating instances where multiple estimates or ranges were reported or estimated for the same location. The yellow curve represents the best-fit to these paired EIR-FOI values (***Smith et al., 2010***). The purple hollow diamond and plus represent the Ghana data, showing our FOI estimates using the two methods and the EIR measured in the field by the entomological team (***Tiedje et al., 2022***).

The data show that mean annual FOI values are consistently below an empirical limit of 20. Due to the highly non-linear relationship between FOI and EIR, there is no single constant factor to convert FOI to EIR (or vice versa) across different settings. In high-transmission regions, transmission efficiency is extremely low: annual EIR can reach a few hundred to a thousand, while annual FOI ranges from about 5 to 10. For future applications, when using our proposed methods to estimate FOI from MOI under sparse sampling schemes, we can rely on this functional curve to convert FOI estimates to corresponding EIR values. There is however an inherently high variance in this conversion. Overall, our paired EIR (measured directly by the entomological team in Ghana (***Tiedje et al., 2022***)) and FOI estimates align well with previous studies, indicating the consistency of our methods.

### The variance inferred by the two-moment approximation method reflects transmission intensity and heterogeneity across individuals

We focused primarily on the mean FOI in both simulation outputs and empirical data. However, the two-moment approximation method also yields estimates of the variance in infection interarrival times. When examining the inferred variance across simulated scenarios, we find that the estimated individual-level mean FOI, when aggregated across the local population, robustly reflects the total number of infections accumulated within that population. This relation holds across a wide range of assumptions, including seasonality, systems openness, heterogeneity in exposure risks, and the distribution of infection interarrival times. In other words, it remains robust regardless of the magnitude of variance in FOI.

The inferred variance is most significantly correlated with the inferred mean FOI. Specifically, a smaller mean FOI is associated with a larger variance (Appendix 1-Figure 26). Overall, seasonal runs exhibit greater variance than non-seasonal runs. Runs with heterogeneous transmission (Appendix 1-Figure 6C) have higher variance compared to homogeneous transmission runs. These findings align with expectations, as both seasonality and transmission heterogeneity increase the dispersion of FOI and inter-arrival times of infections.

## Discussion

Building on estimates of the multiplicity of infection under sparse sampling schemes, we demonstrate the feasibility of converting these values into estimates of the force of infection. In various simulation scenarios using an extended stochastic agent-based model, the two-moment approximation and Little’s Law methods provide good and replicable FOI estimates based on MOI estimates obtained with the Bayesian formulation of the *var*coding method and the bootstrap imputation approach, despite common sampling limitations.

Both methods tend to slightly underestimate FOI due to a bias in underestimating MOI values. The current MOI estimation procedure, specifically the Bayesian formulation of the *var*coding method, does not correct for the limited overlap of *var* genes between co-infecting strains. This limited overlap reduces the number of *var* genes identified per individual, while the Bayesian formulation implicitly assumes that each co-infecting strain contributes a unique set of *var* genes, thereby introducing a downward bias in the MOI estimates.

Studies have shown parasite co-transmission from single mosquito bites in high-transmission regions (***Nkhoma et al., 2020***; ***Wong et al., 2017***), predominantly based on clinical or symptomatic infections (***Andolina et al., 2021***; ***Lindblade et al., 2013***), although asymptomatic cases constitute the majority of the malaria transmission reservoir in these regions. Co-transmitted recombinant parasites, more closely related to each other than parasites from different bites, further reduce the number of identified *var* genes (***Wong et al., 2022***; ***Nkhoma et al., 2020***; ***Wong et al., 2017***) per individual. Low or variable parasite densities due to factors like small blood volumes, genomic DNA quality, clinical status, and within host dynamics also affect MOI estimation (***Peyerl-Hoffmann et al., 2001***; ***Bruce et al., 2000***; ***Okell et al., 2012***; ***Farnert et al., 1997***; ***Färnert et al., 2008***; ***Barry et al., 2021***; ***Hergott et al., 2024***). These issues are common to all measures of MOI and direct measures of FOI. We did not correct for these factors when estimating MOI and FOI for the Ghana surveys. In the future, the Bayesian formulation could be extended to account for these confounders in estimating the number of co-infecting strains.

Our proposed methods leverage infection duration data from malaria therapy patients with neurosyphilis who had no prior malaria exposure, making them well-suited for FOI inference in highly vulnerable subpopulations with similarly naive immune profiles. For our Ghana surveys, we focus on children between 1 and 5 years of age, as their immune profiles are closer to those of naive patients than those of older individuals. In the pre-IRS phase of the Ghana surveys, an estimated mean FOI of about 5 per host per year suggests that a 4-year-old child would have experienced around 20 infections, which makes them appear far from naive. However, the extreme documented diversity of *var* genes (***Tiedje et al., 2025***) means that even with 20 infections, a 4-yearold may have developed immunity to only a small fraction of the total antigenic diversity encoded by these genes. Consequently, they are not as immunologically experienced as it might initially seem. Moreover, studies have shown that long-lived infections in older children and adults can persist for months or even years, including during the dry season. This persistence is driven by high antigenic variation of *var* genes and associated incomplete immunity. Additionally, parasites can skew PfEMP1 expression to produce less adhesive erythrocytes, enhancing splenic clearance, reducing virulence, and maintaining extended periods of sub-clinical parasitemia (***Andrade et al., 2024***; ***Tran et al., 2013***; ***Zhang and Deitsch, 2022***). The impact of immunity on infection duration with age for falciparum malaria remains a challenging open question.

We recognize the limitations that this aspect of infection duration introduces in the FOI estimation. To reduce mis-specification in infection duration and fully utilize our proposed methods, future data collection and sampling could prioritize subpopulations with minimal prior infection and an immune profile similar to that of naïve adults, such as in infants and toddlers. As these individuals are also the most vulnerable, prioritizing them aligns with the short-term priority of all intervention efforts: to monitor and protect the most vulnerable individuals from severe symptoms and death.

The application of both methods generally requires a time series observed over a long period or many realizations of the same series at a single time. However, empirical surveys often rely on sparse sampling schemes with single-time-point observations per host available. We therefore use the population-level queue length distribution by aggregating MOI estimates across all sampled individuals to approximate the steady-state queue length distribution, without explicitly considering individual heterogeneity due to transmission. The estimated FOI, combined with demographic information on population size, provides an estimate of the total number of *Plasmodium falciparum* infections acquired by all hosts in the population per year. We evaluated the impact of individual heterogeneity due to transmission on FOI inference using simulations. Even for significant heterogeneity among individuals, our methods show performance comparable to that of homogeneous scenarios. Additionally, our methods perform similarly for both non-seasonal and seasonal transmission scenarios. Alternative frameworks, such as fitting a non-homogeneous Poisson process or a Gamma process to derive time-varying average arrival rates, or jointly estimating arrival rates and infection duration, are not considered due to their requirements for densely sampled time-series data.

After validating our methods with simulations, we applied them to surveys in the Bongo District of northern Ghana, a high-transmission endemic region where estimating MOI and related FOI has been challenging. The three-round transient IRS intervention proved strong and effective, resulting in a significant reduction in FOI of more than 70%.

Our Ghana surveys lack direct FOI measurements, which prevents us from directly evaluating our methods as we did with simulation outputs. Empirical MOI-FOI pairs from cohort studies are still lacking, and direct FOI measurements are prone to errors due to challenges in differentiating new infections from the temporary absence and re-emergence of old infections. These challenges arise from the low resolution of polymorphic markers used in cohort studies and the complexity of within-host dynamics. Alternative approaches fit epidemiological models to densely sampled cross-sectional surveys without direct FOI measurements, leaving no ground-truth FOI values against which to benchmark or validate their estimates. In these other approaches, model parameterization relies on capturing certain epidemiological quantities, such as prevalence or incidence, similar to the one done in this work. We selected FOI values that maximize the likelihood of observing given MOI distributions. Additionally, we paired our estimated FOI value for Ghana surveys with independently measured EIR (***Tiedje et al., 2022***). We demonstrated a reasonable alignment between our paired EIR-FOI values with the general relationship from previous studies. We acknowledge, however that our validation for field data is indirect and further complicated by high variance in the relationship between EIR and FOI from previous studies.

The FOI estimates obtained through these methods go beyond describing basic malaria epidemiology and evaluating intervention outcomes. FOI for naïve hosts is a fundamental parameter for epidemiological models. The FOI of non-naïve hosts is typically a function of their immune status, body size, and the FOI of naïve hosts. Additionally, the FOI estimates can inform process-based models for the population dynamics of complex infectious diseases, serving as priors for parameterizing and validating more complex agent-based or equation-based models, in a way that reduces computational cost, improves efficiency, and minimizes identifiability issues.

A key characteristic of malaria transmission is the saturation in FOI at high transmission, and the highly non-linear relationship between FOI and EIR. While EIR can reach values of several hundred to a thousand per year, annual FOI typically saturates below 20 (***Smith et al., 2010***). Different choices of field measures of EIR and FOI cannot account for this drastic difference in magnitude. Transmission is highly inefficient in high-transmission regions with high annual EIRs. The difference between these two quantities is mediated primarily by immunity, or within-host dynamics, measurement bias, and heterogeneous transmission (***Donovan et al., 2007***; ***Macdonald, 1950***; ***John et al., 2005***; ***Doolan and Martinez-Alier, 2006***). Mathematical models commonly use the probability of transmission from an infectious mosquito bite to bridge FOI and EIR, as a general parameter encapsulating a variety of processes.

FOI saturation poses significant challenges to intervention efforts in high-transmission endemic regions. In these areas, intervention efforts must dramatically reduce EIR by several orders of magnitude to bring FOI below saturation levels. In other words, high-coverage interventions are needed to achieve any noticeable impact on individual exposure risk. Theoretical models suggest a sharp non-linear transition towards sustainable low transmission or elimination, influenced by the high antigenic diversity of *P. falciparum* (***de Roos et al., 2023***; ***Zhan et al., 2024***). The same molecular information used here to estimate MOI and FOI in the Bongo District underlies estimates of this diversity. Recent ABM modeling on resurgence dynamics indicates that highly resilient systems can compensate for reduced FOI following intervention through heterogeneity in infection duration and selection for longer-lasting infections (***Zhan et al., 2024***). Consequently, high prevalence and MOI levels can persist during interventions in the regime below FOI saturation.

The proposed methods are applicable to evaluate transmission intensity in pathogens exhibiting multi-genomic infection due to large antigenic diversity, including those encoding such variation with multigene families (***Deitsch et al., 2009***). Easier estimation of changes in transmission intensity should enhance the efficiency and evaluation of control programs across a broader range of infectious diseases.

## Materials and Methods

### High genetic diversity of *var* and the associated strain structure of limiting similarity

We briefly describe the biology of the malaria parasite *P. falciparum* that underpins our MOI estimation procedure, the Bayesian formulation of the *var*coding method.

In high-transmission endemic regions, human hosts remain susceptible to malaria re-infection throughout their lifetime (***Doolan et al., 2009***). High asymptomatic prevalence and high MOI result from high transmission rates and incomplete host immunity due to the parasite’s high antigenic variation (***Deitsch et al., 2009***). Parasites achieve this variation and evade the immune system by encoding key variant surface antigens (VSAs) using multigene families (***Deitsch et al., 2009***). One important multigene family in the malaria parasite *P. falciparum* is known as *var*, which encodes PfEMP1 (*Plasmodium falciparum* erythrocyte membrane protein 1), the major VSA during the blood stage of infection (***Zhang and Deitsch, 2022***; ***Baruch et al., 1995***; ***Smith et al., 1995***; ***Su et al., 1995***). Each parasite carries 50-60 *var* genes across its chromosomes, encoding different variants of this protein, which are expressed largely sequentially (Appendix 1–Simulation data, subsection “An extended *var* model,” sub-subsection “Within-host dynamics”).

Empirical sequencing of *var* genes focuses on the DBL*α* tag, a conserved ∼450bp region encoding the immunogenic Duffy-binding-like alpha domain of PfEMP1 (***Tiedje et al., 2025***; ***Ruybal-Pesántez et al., 2017, 2022***; ***Day et al., 2017***). Bioinformatic analyses of a large database of exon 1 sequences of *var* genes revealed a predominantly 1-to-1 DBL*α*-*var* relationship, meaning each DBL*α* tag typically represents a unique *var* gene (***Tan et al., 2023***). Hereafter, we use DBL*α* types and *var* genes interchangeably.

In high-transmission endemic regions, local parasite populations exhibit a vast pool of *var* gene variants, ranging from thousands to tens of thousands (***Day et al., 2017***; ***Tiedje et al., 2022***). These variants are generated primarily through mitotic recombination, but also through meiotic recombination, mutation, and host/mosquito vector migration (***Claessens et al., 2014***; ***Frank et al., 2008***; ***Freitas-Junior et al., 2000***; ***Bopp et al., 2013***). This large pool, combined with negative frequencydependent selection (NFDS) mediated by hosts’ specific immunity, results in the limited overlap of *var* genes among individual repertoires (individual parasite genomes) and isolates (sets of individual parasite genomes co-infecting individual hosts) (***Day et al., 2017***; ***He et al., 2018***). Major groups of *var* genes are classified based on their 5’-flanking region, called ups, which controls gene expression: upsA and upsB/C (non-upsA) (***Rask et al., 2010a***). Non-upsA DBL*α* sequences are ∼ 20 times more diverse and less conserved among repertoires than the upsA DBL*α* sequences. Hence our MOI estimation leverages non-upsA DBL*α* types, as detailed in the following section.

### Bayesian formulation of the “*var*coding” method for MOI estimation

The limited overlap of *var* repertoires allows MOI estimation based on the number of non-upsA DBL*α* types identified from an isolate. The original *var*coding method assumes a constant repertoire length, i.e., number of non-upsA DBL*α* types in a parasite genome, to convert the number of types identified in an isolate to the estimated MOI. This method does not account for the measurement error (Appendix 1-Figure 6D) in this length introduced by the under-sampling or imperfect detection of *var* genes in an infection. We recently extended this method to a Bayesian formulation that considers this error and provides a posterior distribution of MOI values for each sampled individual (***Tiedje et al., 2025***). We documented the steps of this Bayesian formulation, compared two ways of obtaining population-level MOI distribution (either pooling the maximum a posteriori MOI estimates or calculating a mixture distribution), and examined the impact of different priors (***Tiedje et al., 2025***). In our analyses here, we provide the estimated population-level MOI distribution obtained from a mixture distribution using a uniform prior for individuals.

### Empirical surveys from Ghana

We use empirical data from an interrupted time-series study conducted in Bongo District, northern Ghana. This study involves four age-stratified cross-sectional surveys of ∼2,000 participants each, conducted between 2012 and 2016. The study assessed the impacts of a transient three-round IRS intervention, combined with long-lasting insecticidal nets (LLINs), on the asymptomatic *P. falciparum* reservoir (***Tiedje et al., 2017, 2022, 2025***). Surveys were conducted at the end of the wet/high-transmission season (i.e., October) or the dry/low-transmission season (i.e., May/June). The study consists of two phases: (1) Pre-IRS: two surveys before the IRS intervention (Survey 1 in October 2012; Survey 2 in May/June 2013); and (2) Immediately post-IRS: two surveys immediately following the three-round IRS intervention (Survey 3 in October 2015; Survey 4 in May/June 2016) (Appendix 1-Figure 6E). Details on the study area/population, study design, malaria control interventions (IRS and LLINs), inclusion/exclusion criteria, data collection/generation procedures, and routine surveys of antimalarial treatment usage in a previous window of time have been previously described (***Tiedje et al., 2017, 2022, 2025***).

### The under-sampling of infections in empirical surveys

The empirical MOI estimates in various epidemiological studies, including ours in Bongo District, northern Ghana, rely on microscopy-positive individuals (***Tiedje et al., 2025, 2022***). Due to microscopy’s limited sensitivity, a significant fraction of individuals who carry infections are undetected. A subset of Ghana surveys also include submicroscopic infections detected by PCR (***Tiedje et al., 2025, 2022***), which is significantly more sensitive and can detect a higher fraction, if not 100%, of individuals with *P. falciparum* infections. Using surveys with both microscopy and PCR detection, we estimate conversion factors of 0.76 for untreated children and 0.67 for antimalarial-treated children aged 1–5 years.

For surveys with both detection methods, we directly calculate the number of microscopy-negative but PCR-positive individuals. For surveys with only microscopy data, we use the estimated conversion factors to estimate the number of microscopy-negative but PCR-positive individuals. Additionally, we account for the high but not exactly known sensitivity of PCR by assuming its detectability ranges from relatively low (10% of all PCR-negative individuals carrying undetected infections) to perfect (none of PCR-negative individuals carrying undetected infections). We calculate the number of individuals with undetected infections by PCR for each sensitivity level.

After estimating the number of individuals with undetected infections (both microscopy-negative but PCR-positive and PCR-negative but infected), we sample from existing MOI estimates of microscopy-positive individuals not under antimalarial treatment (see the following section) to represent the missing MOI data.

Similarly, for individuals who are microscopy-positive but lacking *var* information due to factors like low DNA quality, we sample values from existing MOI estimates of microscopy-positive individuals not under antimalarial treatment to represent the missing MOI data.

We assume that microscopy-negative but PCR-positive children aged 1-5 years and microscopy-positive children aged 1-5 years have similar MOI distributions. This assumption is suggested by our analysis of Ghana surveys, which shows no clear relationship between parasitemia levels and MOI (or the number of *var* genes detected within an individual host, on the basis of which our MOI values were estimated) (Appendix 1-Figure 7). We scale the parasitemia levels and the number of non-ups A *var* genes or MOI estimates before performing the regression. Parasitemia levels underlie the difference in detection sensitivity between PCR and microscopy.

This lack of a clear relationship can be attributed to several factors. One factor is immune regulation of parasite density, where host immunity may limit parasite density without reducing the diversity of co-infecting strains (***Eldh et al., 2019***), leading to individuals with low parasitemia but high MOI. Another factor is asynchronous parasite dynamics, where different parasite clones replicate asynchronously (***Färnert et al., 1997***), resulting in varied parasite densities that do not directly correlate with the number of distinct strains present. This could explain why individuals with low parasitemia still exhibit multiple strains. Lastly, competition among parasite strains suppress the growth of individual clones, lowering parasite densities while maintaining high strain diversity, thus reducing the expected correlation between MOI and parasitemia (***Sondo et al., 2019***; ***Earland et al., 2019b***).

### Antimalarial drug treatment of infections in empirical surveys

Individuals may seek and receive antimalarial treatment in response to symptoms or perceived transmission risk. In our surveys from northern Ghana, over 50% of children aged 1-5 years responded that they had received an antimalarial treatment in the previous two-weeks (i.e., participants that reported they were sick, sought treatment, and were provided with an antimalarial treatment) prior to the wet/high-transmission survey before IRS (i.e., Survey 1, Appendix 1-Figure 6E) (***Tiedje et al., 2022***). This fraction is significantly lower for the dry/low-transmission survey before IRS and the surveys collected immediately after IRS (i.e., Survey 2-4, Appendix 1-Figure 6E) (***Tiedje et al., 2022***).

Disentangling the effect of drug treatment on measurements like infection duration is challenging. Since our methods use infection duration data from naïve malaria therapy patients with neurosyphilis, drug treatment can potentially violate this assumption. We propose two solutions: (1) exclude treated individuals from the analysis; (2) remove treated individuals’ samples and use a bootstrap imputation approach based on the remaining population. Specifically, we sample from the MOI estimates of untreated microscopy-positive individuals to represent MOI estimates for treated individuals, which corrects for individuals who have used antimalarial drugs and show either no infection (MOI = 0) or infection (MOI > 0). Hence, this solution provides an upper bound for FOI estimates. Numerical simulations show our bootstrap imputation approach is robust even with a significant fraction of treated individuals, as seen in our Ghana surveys.

The final distribution of MOI estimates at the population level includes values for microscopy-positive individuals, imputed values for individuals with missing MOI information or false negatives, imputed values for treated individuals (for the second solution), and true zeros for uninfected individuals. This distribution is used for FOI inference.

### Inferring FOI from MOI estimates

#### Malaria transmission in relation to queuing theory

In a cohort of individuals acquiring and clearing infections independently, infections occurs as a homogeneous Poisson process with a rate equal to the mean FOI, and each infection has an exponentially distributed duration. At equilibrium, MOI follows a Poisson distribution with a mean equal to the mean FOI divided by the mean clearance rate (***Dietz et al., 1974***). In practice, individuals are often capacity-limited, such that they can only carry up to a certain number of concurrent infections due to within-host competition or immune regulation. New infections that arrive when a host is already at capacity can be simply blocked rather than queued or delayed, resulting in an MOI that follows a Poisson distribution truncated at the carrying capacity and normalized. However, infection arrivals frequently deviate from a homogeneous Poisson process because of factors such as seasonality and heterogeneity in exposure risk, and infection duration may also deviates from an exponential distribution. Consequently, the MOI distribution is often overdispersed. In this case, no simple analytical relationship exists between the MOI distribution and the mean FOI or clearance rate. We formally test for deviations from Poisson homogeneity against a negative binomial alternative in both simulated and empirical MOI distributions, as detailed in Appendix 1-Test of deviation from Poisson homogeneity in MOI estimates and supplementary file 2-deviationFromPoissonTest.xlsx. Notably, infection durations observed in malaria-naive patients from historical neurosyphilis treatment studies, where patients were intentionally infected with malaria, also exhibit a standard deviation substantially different from their mean (***Collins and Jeffery, 1999***; ***Maire et al., 2006***) (see details below).

Despite these complexities, the qualitative relationship between MOI and FOI is intuitive when information on infection duration is known. Higher FOI values should correspond to higher MOI values. Less variable FOI values should result in narrower or more concentrated MOI distributions, whereas more variable FOI values should lead to more spread-out MOI distributions.

The process of acquiring infectious bites is structurally analogous to stochastic queuing theory, which relates queue length to the intensity of arrivals, priority schedules, and service and waiting times. Modeled using differential equations (with the Kolmogorov equations), queueing systems comprise three main components: queue length, the intensity of arrivals, and service times. Knowing any two allows for the inference of the third.

In malaria transmission, hosts resemble service facilities composed of a collection of servers, with each infection akin to a customer. Just as service facilities have a carrying capacity for the number of customers they can serve simultaneously, hosts have a carrying capacity for blood-stage infections, which limits the maximum number of infections they can harbor (Appendix 1-Figure 8). Empirical MOI estimates provide information on queue length. Hypothetically, knowing the service times, i.e., infection durations, can help infer the intensity of arrivals, i.e., the rate at which hosts acquire infections or FOI.

However, determining infection durations is challenging in endemic areas where multi-genomic infections are common. Popular polymorphic markers often fail to distinguish between co-infecting strains (***Argyropoulos et al., 2023***), complicating the tracking of the emergence and clearance of individual strains from the peripheral blood. Complex within-host dynamics further complicate tracking unless daily sampling is conducted, which is impractical in real settings. Frequent ectopic recombination of *var* genes complicates assigning genes to specific chromosomal locations. This difficulty in phasing compromises the integrity of individual strains, making it hard to isolate them and track their first appearance and subsequent clearance in blood over time. Additionally, infection duration varies widely across age groups, geographical locations, and sampling times (***Childs and Buckee, 2014***; ***Chang et al., 2016***; ***Ashley and White, 2014***; ***Bretscher et al., 2011***).

We therefore propose focusing on FOI inference in subpopulations with naive or near-naive immune profiles. Their infection duration can be approximated by that of naive hosts, as seen in a historical medical study of neurosyphilis patients intentionally infected with malaria as a treatment. Between 1940 and 1963, 318 syphilis patients were infected with a single strain of *P. falciparum* (***Collins and Jeffery, 1999***; ***Maire et al., 2006***), and data on fever and parasite counts in the blood were recorded. Since these patients had no prior *P. falciparum* infections, the documented infection duration reflects that of naive infections.

In Ghana surveys, we focus on children aged 1-5 years, who have accumulated far fewer infections and less immune memory compared to older individuals, an aspect we discuss in the Discussion section. We treat these children as nearly naive and approximate their duration infection with that of naive hosts. Using their MOI estimates, we infer FOI for these children using the following two methods from queuing theory.

#### A two-moment approximation for a queue of finite capacity

Analysis of multi-server models is challenging, with exact results available only for specific cases, such as the previously mentioned *M*/*M*/*c*/*k* models. In these models, *M* represents exponential inter-arrival and service times, *c* is the number of servers, and *k* is the maximum queue capacity, including both customers being served and those waiting. Additional models like *M*/*G*/*c*/*c* and *GI*/*M*/*c*/*c* +*r* queues also require exponential distributions for either inter-arrival or service times, where *G* and *GI* represent generic random variables and independent and identically distributed (i.i.d.) generic random variables, respectively. These models can often be too restrictive for real-world scenarios as well.

We examine a two-moment approximation method introduced by Kim and Cha (***Choi et al., 2005***). This method considers the *GI*/*G*/*c*/*c* + *r* queue, where inter-arrival times (*GI*) and service times (*G*) of customers are independent sequences of i.i.d. general random variables *A* and *S*, respectively. There are *c* (≥ 1) identical servers in parallel and *r* (≥ 0) waiting places. In this framework, overdispersion in MOI arises from temporally structured (non-Poisson) acquisition processes that generate bursts of infection, compounded by non-exponential infection durations, among otherwise homogeneous hosts.

Let *N* denote the number of customers in the system at an arbitrary time, and *N*^*A*^(*N*^*D*^) denotes the number of customers that an arriving customer finds (that a departing customer leaves behind) in steady state. Customers who arrive to find *c* + *r* customers in the system depart immediately, leaving those *c* + *r* customers behind. Let 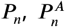 and 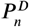 denote the probabilities that *N* = *n, N*^*A*^ = *n*, and *N*^*D*^ = *n*, respectively, for 0 ≤ *n* ≤ *c* + *r*. These probabilities are expressed in terms of the following quantities:

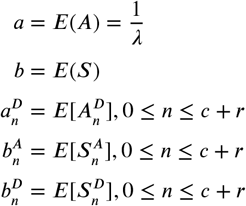

Where 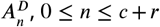, is the residual inter-arrival time at the departure instant of a customer who leaves behind *n* customers in the system, and 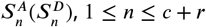, is the residual service time of a randomly chosen busy server at the arrival instant (the departure instant) of a customer who finds (leaves behind) *n* customers in the system. From these definitions, we have 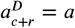 and 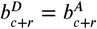. We set 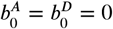. We assume that all the above quantities are well defined and finite.

Using Theorems 4.3.19 and 4.3.43 from Franken et al (***Heyman, 1985***), the steady-state queue-length distribution can be derived:

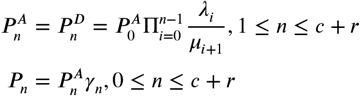

where

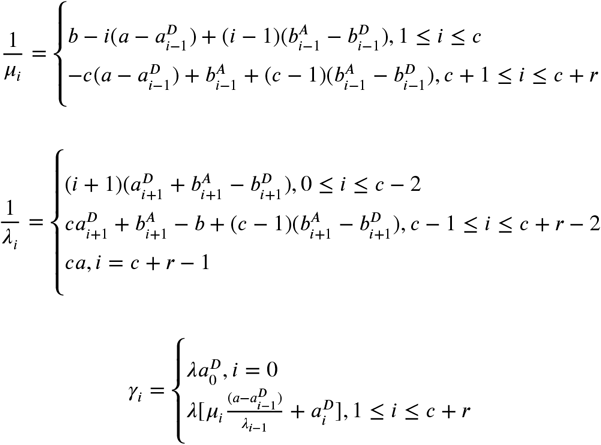

And, by normalization, 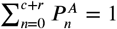:

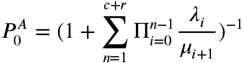

Since quantities 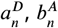, and 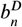 are difficult to compute in general, Kim and Cha propose an approximation to the exact expression which replaces these unknown arrival- and departure-average quantities by their corresponding (well-known) time-average counterparts, which are exact for exponential inter-arrival and service times. That is:

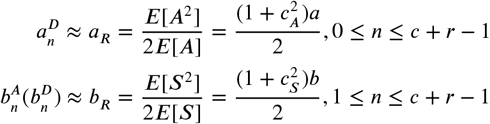

Where 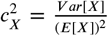 is the square coefficient of variation (SCV) of a random variable *X* with distribution function *F*.

Therefore, a two-moment approximation for the steady-state queue-length distribution is:

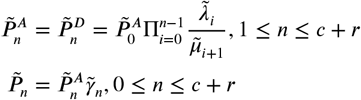

where

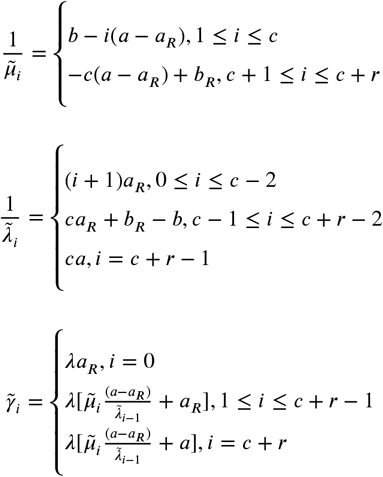

And, by normalization, 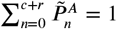:

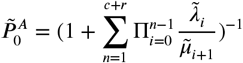

#### Likelihood formulation and parameter estimation

We vary the mean and variance parameters for inter-arrival times across wide ranges (supplementary file 4-meanAndVarianceParams.xlsx). For each mean and variance combination, we calculate the steady-state queue length distribution, i.e., the probability density distribution of MOI, using the two-moment approximation method. The goal is to identify the parameter combination that minimizes the negative log-likelihood (or maximizes the likelihood) of observed MOI distributions from simulated outputs or Ghana surveys:

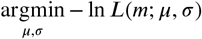

with the likelihood defined as follows:

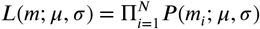

where *N* is the number of individual hosts, *m*_*i*_ is the MOI estimate for individual *i, m* is the vector of MOI estimates for all hosts, *μ* and *σ* represent the mean and variance parameters, respectively, and *P* is the steady state queue length distribution from the two-moment approximation method with specific mean and variance parameter values, as defined in the previous section.

The shape of the negative log likelihood for both simulated outputs and Ghana surveys is concave upwards around the trough, signifying a clear minimum point (Appendix 1-Figure 9). We tested the impact of different grid value choices on the FOI inference results by refining the grid to include more points, ensuring the FOI inference results are consistent. Specifically, we reduce the grid width for the mean parameter to half and a quarter of the original width, and for the variance parameter to half, a quarter, an eighth, and a sixteenth of the original width. The FOI inference results remain either unchanged or within a 1% deviation from those based on the original grid width (Appendix 1-Figure 10).

Details for deriving confidence intervals for the estimated parameters are provided in Appendix 1-Confidence intervals for FOI inference.

#### Choice of *c* and *r* for the *GI*/*G*/*c*/*c* + *r* queue in the two-moment approximation method

When applying the two-moment approximation method, values for the number of parallel servers (*c*) and waiting places (*r*) need to be specified. Since MOI is defined exclusively for blood-stage infections, *r* is set to 0 by default. The parameter *c* corresponds to the carrying capacity of blood-stage infections. The maximum MOI observed in empirical data from Bongo, based on the *var*coding method, is 20. Certain factors which reduce the number of *var* genes identified in an individual, and thus affect MOI estimation, are not explicitly accounted for in the current MOI estimation (see Discussion), so the actual carrying capacity could be higher. For simplicity, we assume the value of *c* to be 30 in the simulation. Provided that *c* is kept consistent across simulations and the two-moment approximation method, this choice should not affect FOI inference. For empirical surveys from Bongo, we set *c* to 25, 30, 40, and 60 to systematically investigate its impact on FOI inference results. The FOI inference results are similar across these values (Figure 3 and Appendix 1-Figure 23-25).

In general, the choice of *c* depends on the maximum MOI observed in a given empirical dataset under high transmission. To account for factors that may lead to underestimation of MOI, *c* should be set higher than the observed maximum MOI. Since Bongo District of northern Ghana is a high-transmission endemic region, we expect the range of its *c* to be applicable to other empirical datasets.

#### The mean arrival rate of infection from Little’s Law

The second method is known as Little’s Law (***Little and Graves, 2008***), which describes a relationship between the three main components of queuing systems. This law states that the average number of items in a queuing system *L* equals the average arrival rate *λ* multiplied by the average waiting time of an item in the system, *W*. Reformulating Little’s Law for malaria transmission, the average arrival rate of infection *λ* equals the average number of blood-stage infections present in an individual *L* divided by the average duration of infection *W*.

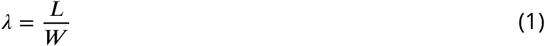

The relationship is simple and general, holding true regardless of the number of servers (carrying capacity of blood-stage infections in hosts), the service time distribution (infection duration distribution), the distribution of inter-arrival times, the order of service, or the queue structure.

#### Population-level MOI distribution for approximating time-series observation of MOI per host or many realizations at the same sampling time per host

Both the two-moment approximation and Little’s Law methods require either the steady-state queue length distribution (MOI distribution) or the moments of this distribution. Obtaining such a steady-state queue length distribution necessitates either densely tracked time-series observations per host or many realizations at the same sampling time per host. However, under sparse sampling schemes, we only have two one-time-point observations per host: one at the end of wet/high-transmission and another at the end of dry/low-transmission. This is typically the case for empirical data, although numerical simulations could circumvent this limitation and generate such output.

Nonetheless, we have a population-level queue length distribution from both simulation outputs and empirical data by aggregating MOI estimates across all sampled individuals. We use this distribution to represent and approximate the steady-state queue length distribution at the individual level, not explicitly considering individual heterogeneity due to transmission. We assess the impact of individual heterogeneity due to transmission on FOI inference using simulation outputs (Appendix 1-Figure 6C). The performance of our methods across various simulated scenarios is reported and discussed in the Results and Discussion sections.

### Conversion between FOI and the Entomological Inoculation Rate, or EIR

As an indirect proxy for transmission intensity, malariologists typically measure EIR by counting the number of infectious bites a human host receives within a fixed time interval (***Shaukat et al., 2010***). EIR is considered a standard metric of malaria transmission. Although both FOI and EIR reflect transmission intensity, FOI directly concerns detectable blood-stage infections, while EIR pertains to human-infectious vector contact rates. FOI is defined as the rate at which a host acquires infections, with the focus specifically on blood-stage strains for the following reason. Only blood-stage infections are detectable in all direct measures of FOI. Quantities used in indirect model-fitting approaches for estimating FOI are also based on or reflect these blood-stage strains/infections. Only these blood-stage strains/infections are transmissible to other individuals, impacting disease dynamics.

Studies comparing annual *P. falciparum* EIR and FOI estimates from age-stratified prevalence in cross-sectional parasitological studies have found significantly different magnitudes for these two quantities (***Smith et al., 2010***). The number of blood-stage infections per infectious bite (FOI/EIR) is referred to as transmission efficiency. Multiple studies indicate that malaria transmission is inefficient in high-intensity settings, and the reasons for this have been debated. Potential causes include heterogeneous biting, immunity or within-host dynamics, and measurement bias.

We utilize a functional curve with empirically derived parameters under the assumption of heterogeneous transmission, describing the highly non-linear relationship between reported EIR-FOI pairs (***Smith et al., 2010***). The functional curve, with FOI *h*, EIR *E*, and the corresponding parameters

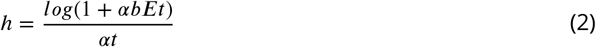

The EIR-FOI pairs (***Smith et al., 2010***) and the functional curve provide a basis for converting between these two quantities. EIR data for a subset of surveys in Bongo District, northern Ghana, were obtained (***Tiedje et al., 2022***). Combined with FOI estimates from our two proposed methods, we generate an EIR-FOI pair for empirical surveys in Bongo District. This enables us to evaluate whether our EIR-FOI pair aligns with historical data and the functional curve with the best-fit parameter values.

## Appendix

### The measurement error

We incorporate a measurement error model (Appendix 1-Figure 6D) into the sampling of simulation outputs and MOI estimation for both simulation outputs and empirical data. This model accounts for the under-sampling or imperfect detection of *var* genes in the field. By sub-sampling *var* genes per strain, we reduce the number of distinct *var* genes available per host. This model is based on the repertoire size distribution, derived from molecular sequences of infections expected to be monoclonal (MOI = 1), i.e., hosts infected by a single *P. falciparum* strain as they had 45 or fewer non-upsA DBL*α* types. These molecular sequences were collected during six cross-sectional surveys conducted from 2012 to 2016 in Bongo District, northern Ghana (Appendix 1-Figure 6E) (***Tiedje et al., 2025, 2022***; ***Pilosof et al., 2019***).

**Figure 5.**
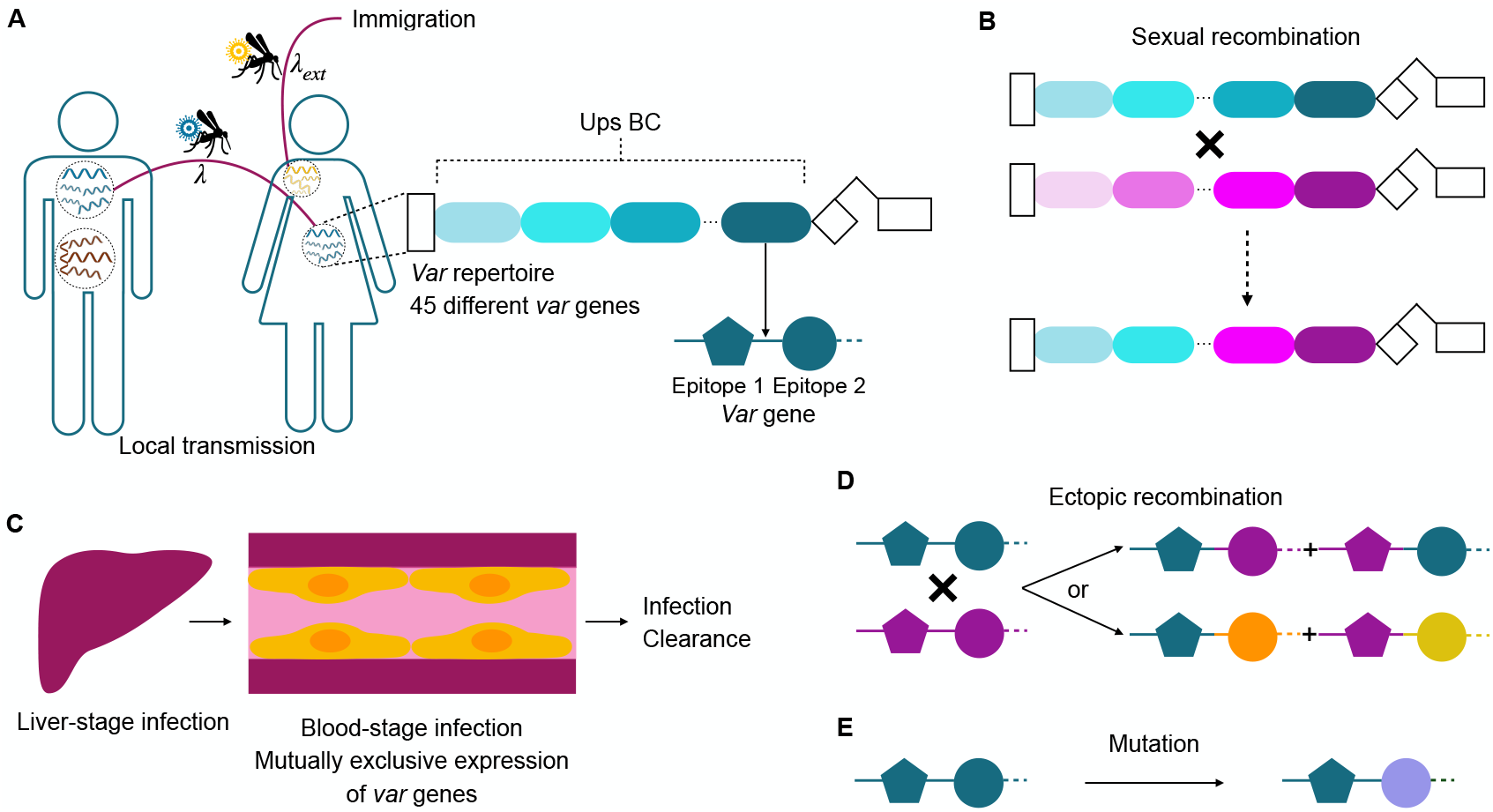
Agent-based model for falciparum malaria transmission. (A) The stochastic model tracks infection history and specific immune memory of individual hosts to variant surface antigens encoded by *var* genes. At transmission events, a donor and a recipient host are randomly selected. Transmission occurs if the donor host has blood-stage infections and the recipient host has not reached carrying capacity of infections in its liver. Each parasite genome in the donor host is transmitted to a mosquito with a probability of 1/(number of genomes) multiplied by the transmissibility of the currently expressed gene. Each parasite genome carries 45 *var* genes, with each gene represented by a linear combination of two epitopes (depicted by different shapes), with many possible variants each (alleles, depicted by different colors). (B) During the sexual stage within mosquitoes, different parasite genomes can exchange *var* genes through meiotic recombination, generating novel recombinant repertoires. The recipient host can receive either recombinant genomes or original genomes. (C) When a repertoire is successfully transmitted to a recipient host, it undergoes a 7-day dormant liver stage before entering the blood stage, where *var* genes are sequentially expressed. If the host has no immunity against either epitope of a given *var* gene, its expression lasts 7 days (and either 7.5 or 8 days in additional simulations). Immunity to one of the two epitopes reduces the expression by approximately half, while complete immunity to both epitopes leads to immediate clearance of the gene product. An infection ends either when all *var* genes in the repertoire have been expressed or recognized, or, alternatively, with a certain probability, before the full repertoire is exhausted (Appendix 1–Simulation data, subsection “An extended *var* model,” sub-subsection “Within-host dynamics”). (D) During the asexual blood stage of infection, *var* genes within the same genome can swap their two epitope alleles through mitotic (ectopic) recombination, generating new epitopes with a certain probability. (E) *Var* genes can also mutate their epitopes to create new genes.

**Figure 6.**
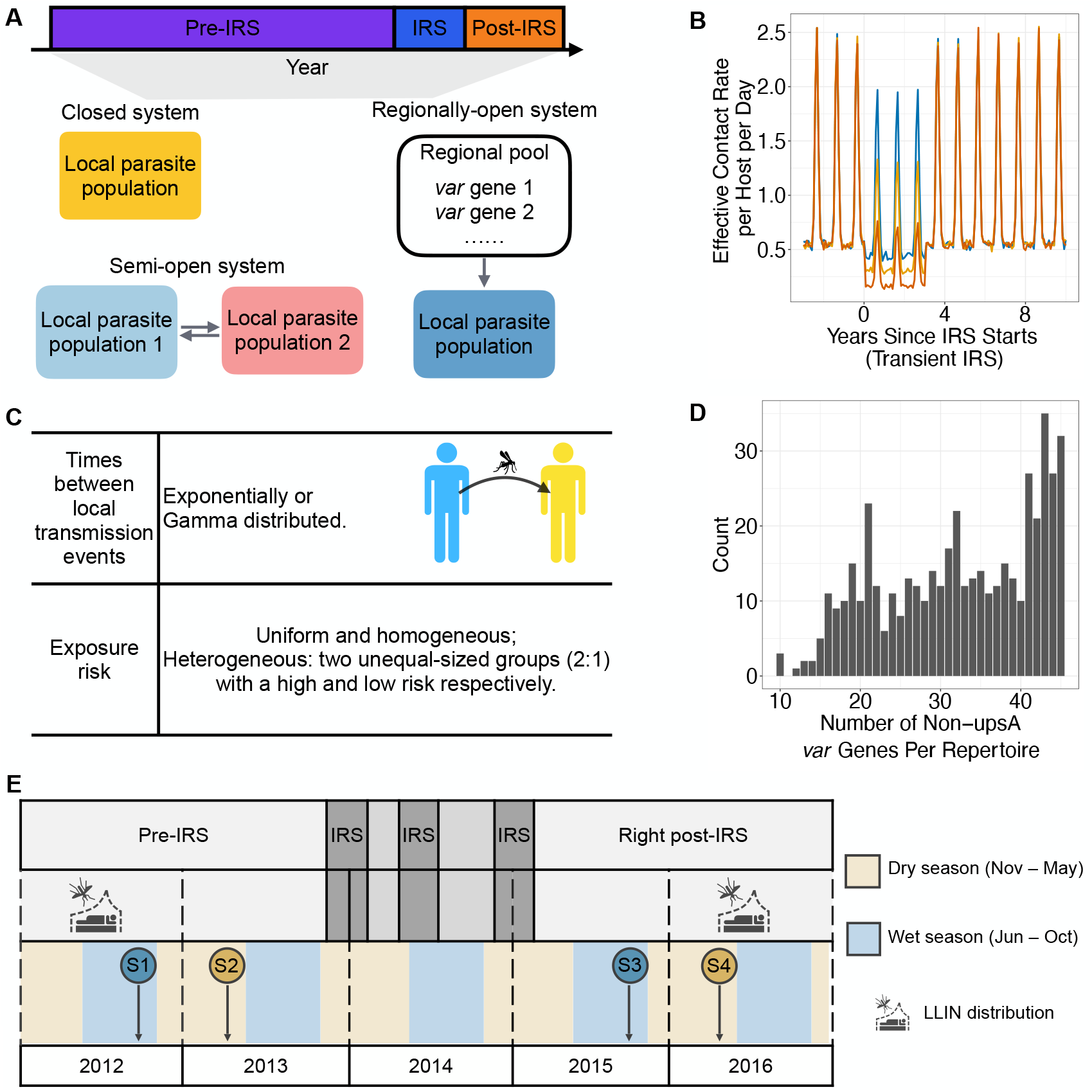
(A) Each simulation comprises three stages: a “pre-IRS” period where local transmission reaches a semi-stationary state, followed by a three-year “IRS” intervention period (transient IRS) which reduces transmission rate, and a “post-IRS” period where transmission rates return to original levels. After transmission initialization, closed systems do not receive migrant genomes from the regional pool. Semi-open systems explicitly model two local populations connected by migration. Regionally-open systems continually receive migrant genomes from the regional pool throughout the simulation. This figure was adapted from [(***Zhan et al., 2024***), Figure 1](CC BY 4.0 license). The copyright holder has granted permission to publish under a CC BY 4.0 license. (B) Transmission intensity or effective contact rate, varies seasonally across the pre-, during-, and post-intervention periods. We simulate three levels of perturbation corresponding to approximately 20% (low-coverage IRS), 40-45% (mid-coverage IRS), and 65-75% (high-coverage IRS) reductions in transmission. Under non-seasonal transmission, the transmission intensity remains constant throughout the year, decreases only during IRS, and then returns to its original level once IRS ends. (C) We examine different statistical distributions for times between local transmission events: exponential and Gamma. We consider homogeneous and heterogeneous exposure risks. In the latter, 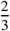 of the population are high-risk, receiving approximately 94% of all bites, while the remaining population receives the rest. (D) The measurement error is depicted as a histogram showing the number of non-upsA (i.e., upsB and upsC) DBL*α* types per repertoire from putatively “monoclonal” infections, characterized by having 45 or fewer non-upsA DBL*α* types. These sequences were collected during six cross-sectional surveys conducted from 2012 to 2016 in Bongo District. This measurement error represents under-sampling or imperfect detection of *var* genes. (E) The study consists of four age-stratified cross-sectional surveys in Bongo District, Ghana, conducted at the end of wet/high-transmission seasons (blue circles) and dry/low-transmission seasons (gold circles). Two phases are covered: (1) Pre-IRS: Survey 1 (S1) in October 2012 and Survey 2 (S2) in May/June 2013; (2) Right post-IRS: Survey 3 (S3) in October 2015 and Survey 4 (S4) in May/June 2016. IRS was implemented with widespread LLIN usage distributed between 2010-2012 and again in 2016 (***Tiedje et al., 2025***; ***Gogue et al., 2020***). This figure was adapted from [(***Tiedje et al., 2022***), Figure 1] (CC BY 4.0 license). The copyright holder has granted permission to publish under a CC BY 4.0 license.

**Figure 7.**
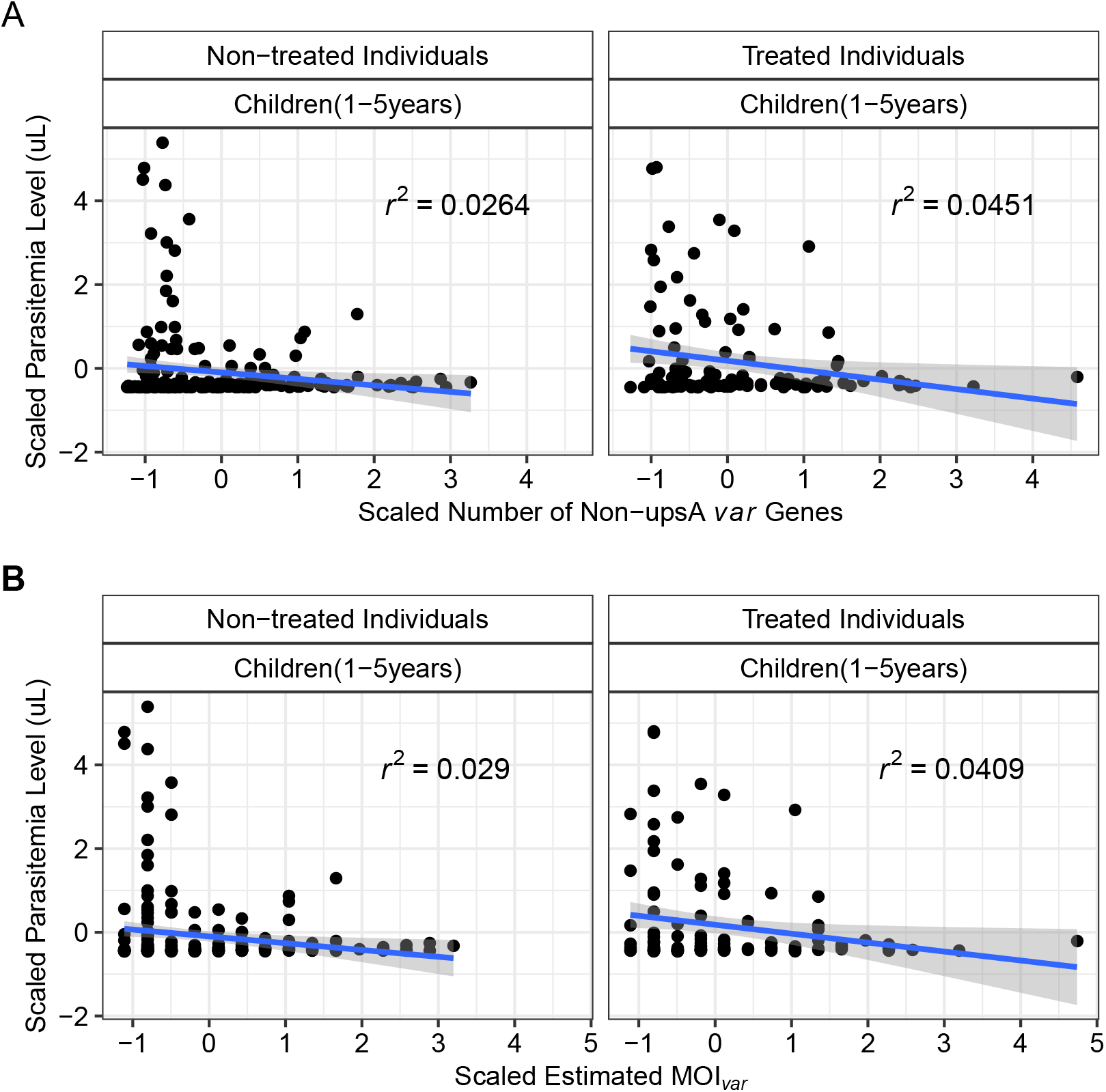
The relationship between the parasitemia level of the individual (measured in uL) and (A) the number of non-upsA *var* genes per isolate/individual, or (B) MOI estimates from the Bayesian formulation of the *var*coding method. There is a lack of association between the x-axis and y-axis variables among both untreated and antimalarial drug-treated individuals. We scale the parasitemia levels and the number of non-ups A *var* genes or MOI estimates before performing the regression.

**Figure 8.**
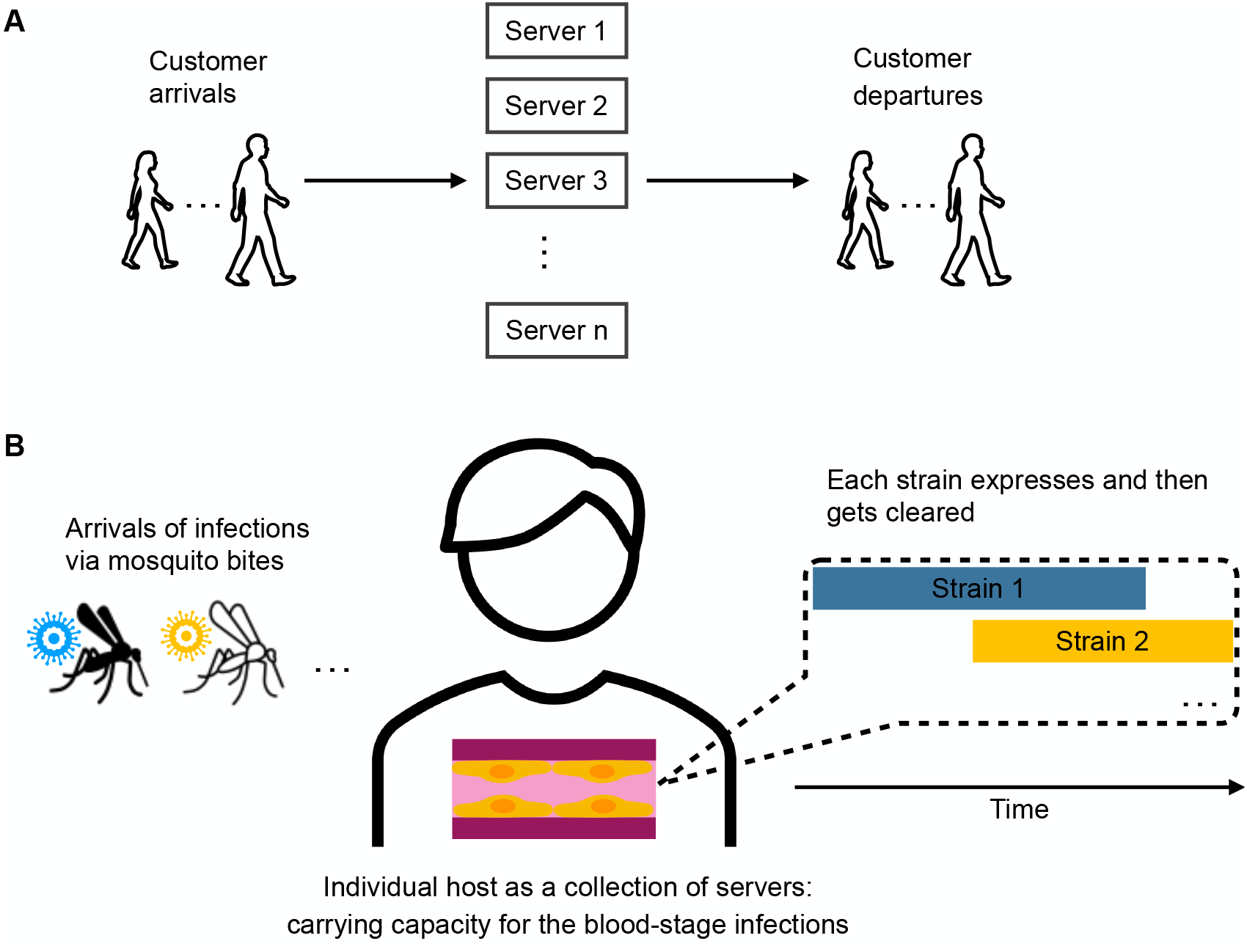
Schematic illustration of (A) systems in queuing theory and (B) malaria transmission.

**Figure 9.**
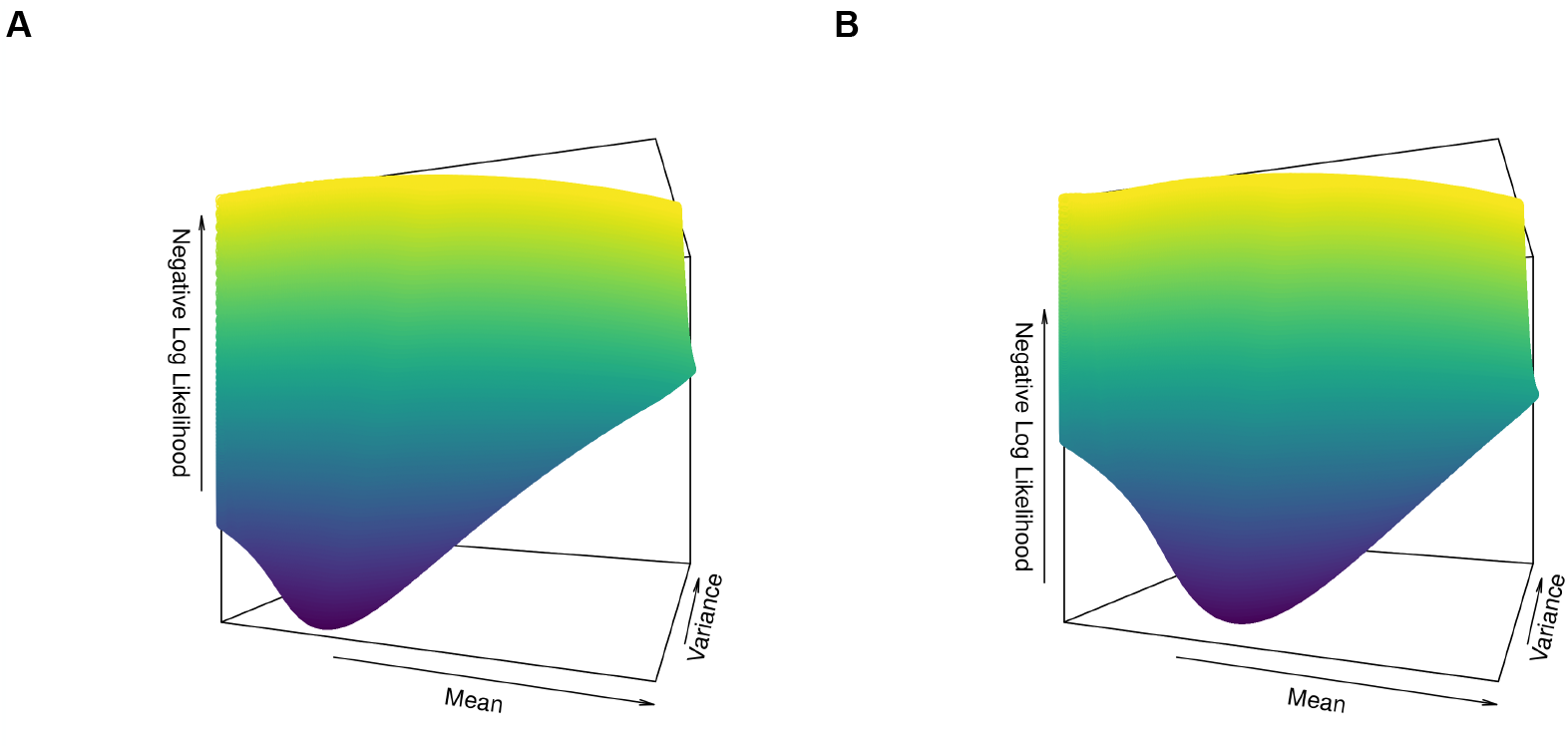
The shape of the negative log likelihood for (A) a simulation run (pre-IRS) with Gamma-distributed times between local transmission events in a seasonal, semi-open system with heterogeneous exposure risk, and (B) Ghana pre-IRS surveys (Survey 1 and 2) with *c* = 30 and mid PCR detectability. We remove the infinite and extremely large values of the negative log likelihood, and plot the rest to improve visualization.

**Figure 10.**
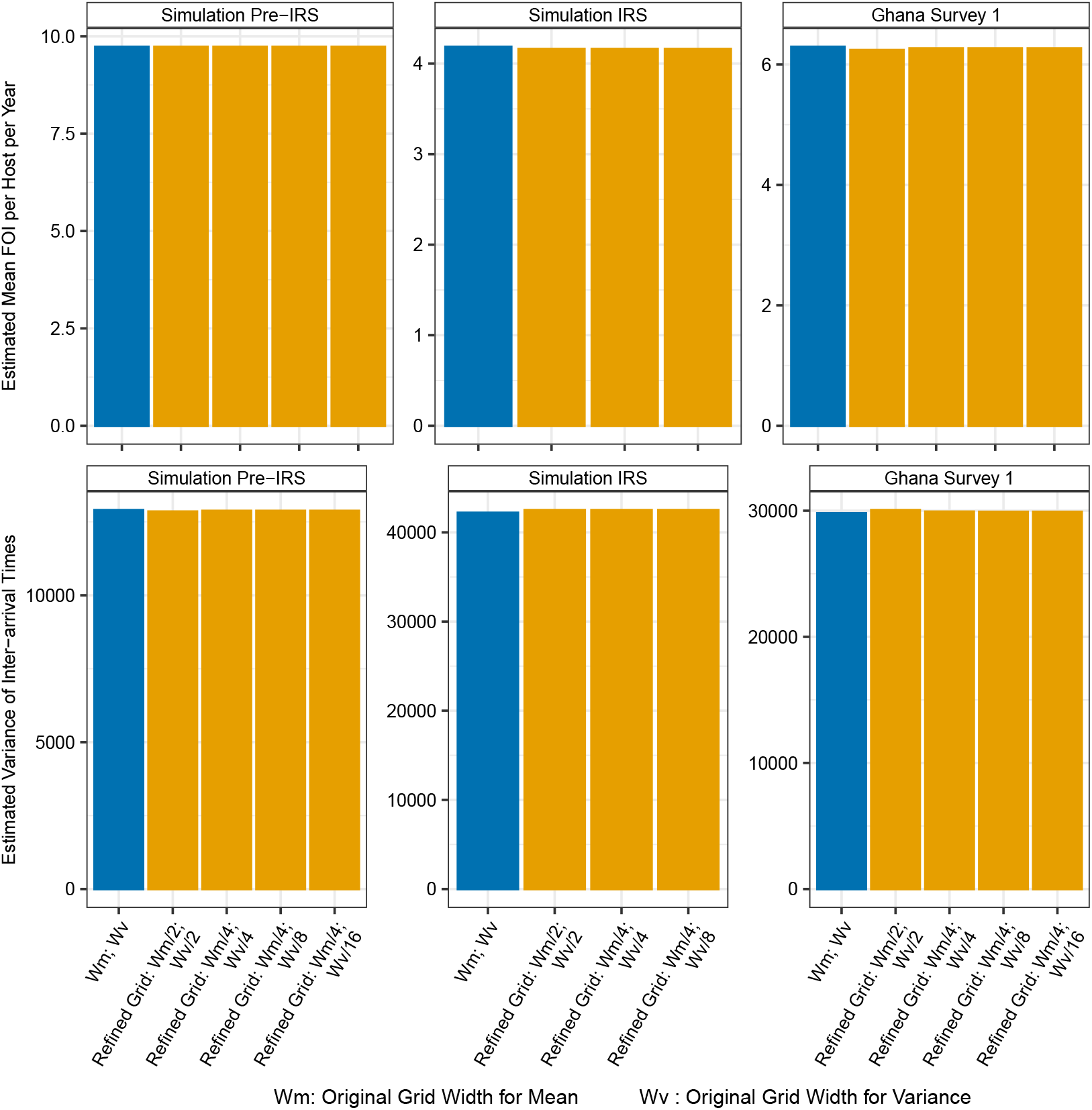
The impact of grid value choices on the results of FOI inference in either simulated outputs or Ghana data. By further reducing the grid width to include more combinations of the mean and variance values of inter-arrival times, the FOI inference results remain either unchanged or deviate by no more than 1% from those based on the original grid width.

## Simulation data

### An extended *var* model

#### Overview

To evaluate the performance of the two proposed queuing theory methods for FOI inference across different transmission settings, we use a computational model, (***He et al., 2018***; ***Zhan et al., 2024***) detailed in this section and illustrated in Appendix 1-Figure 5. This model is an agent-based (individual-based), discrete-event, and continuous-time stochastic model in which all known possible future events are stored in a single event queue along with their putative times, which may be fixed or drawn from a probability distribution with a certain rate. Event rates are chosen based on malaria epidemiology literature, field studies, and in vitro/in vivo values. When an event occurs, it can cause the addition or removal of future events in the queue or the modification of their rates, resulting in a recalculation of putative times. This approach is implemented using the next-reaction method (***Gibson and Bruck, 2000***), an optimization of the Gillespie first-reaction method (***Gillespie, 1976***).

Individual human hosts die and are immediately replaced with newborns who have no immunity. The age structure of the human host population follows a truncated exponential distribution with a mean age of 30 years and a maximum age of 80 years. Individual infections and the immune history of each host are tracked. Evolutionary mechanisms, such as mitotic/ectopic recombination and mutation, are explicitly modeled.

At the beginning of each simulation, a small number of hosts are randomly selected and infected with distinct parasite genomes, assembled from a pool of *var* genes, to initiate local transmission. Mosquito vectors are not explicitly represented as agents in the model; instead, we consider an effective contact rate (referred to as the transmission rate, which under some assumptions is effectively equivalent to vectorial capacity), which determines the times of local transmission events. At these times, a donor and a recipient host are randomly selected, and successful transmission occurs only if the donor has blood-stage infections and the liver stage of the recipient is below the specified carrying capacity. The times between transmission events can follow various distributions, such as exponential and Gamma.

#### *Var* repertoire and gene structure

We assume a parasite repertoire size of 45, based on the median number of non-upsA DBL*α* sequences identified in our 3D7 laboratory isolate (***Ruybal-Pesántez et al., 2022***; ***Tiedje et al., 2022***). The classification of *var* genes into upsA and non-upsA (upsB and upsC) types is based on their semi-conserved upstream promoter sequences (ups) (***Gardner et al., 2002***; ***Lavstsen et al., 2003***; ***Kraemer et al., 2007***; ***Rask et al., 2010b***). Although each parasite carries both types of *var* genes in a fairly constant proportion (***Ruybal-Pesántez et al., 2017***; ***Tiedje et al., 2025***; ***Ruybal-Pesántez et al., 2022***; ***Buckee and Recker, 2012***; ***Rask et al., 2010b***), we focus on the more diverse and less conserved non-upsA sequences for MOI estimation. Despite functional differences, the groupings do not necessarily correlate with function, as there is within-group functional heterogeneity and cross-group functional similarity (***Claessens et al., 2012***; ***Kaestli et al., 2006***; ***Rottmann et al., 2006***). The mechanism behind the fairly constant proportion of these groups in empirical samples across different times and locations remains unclear, so we simplify by considering only the non-upsA type.

Each gene is modeled as a linear combination of two epitopes (alleles), based on the empirical description of the two hypervariable subregions in the *var* tag region amplified from field isolates (***Larremore et al., 2013***).

#### Ectopic recombination

We model ectopic recombination among genes within the same genome during the asexual stage inside the human host. This process is a major mechanism of *var* gene diversification, occurring during both sexual and asexual stages (***Claessens et al., 2014***). For simplicity, we focus on the asexual stage, where two randomly selected genes from a strain undergo recombination. The breakpoint location is chosen randomly. Under normal recombination, alleles of the two genes are swapped with a probability of creating new alleles. Under conversion, the second gene remains unchanged. In this implementation, we assume all events result in normal recombination rather than gene conversion.

Newly recombined genes have a probability of being functional (i.e., viable), influenced by the similarity of their parental genes and the breakpoint locations (***Drummond et al., 2005***):

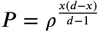

Where *ρ* represents recombination tolerance, *d* is the the genetic distance between the two parental genes, and *x* is the genetic distance between the offspring gene and one of the two parental genes.

Non-functional offspring genes replace their parental ones. As they do not express and get deactivated immediately, they shorten the infection duration of the strain they constitute, thereby reducing its fitness.

#### Meiotic recombination

Meiotic recombination occurs between strains during sexual replication inside the mosquito vector. While we do not explicitly model mosquitoes, we represent meiotic recombination between genomes at the time of a transmission event.

When multiple strains, denoted by *m* (*m* > 1), co-infect a donor host, a Bernoulli trial is conducted for each strain to determine if it will be transmitted via the contact event, with the success probability equal to its transmissibility. Each strain’s transmissibility depends on the currently expressed gene’s transmissibility. Co-infection reduces the transmissibility of each strain by a factor of *m*. Only a subset *n* of these strains, where *n* ≤ *m*, is selected. In nature, this subset would co-infect a mosquito vector.

To simulate meiotic recombination, each of the *n* strains which are to be transmitted from the mosquito vector to a recipient human host is obtained by drawing two parental strains from the pool of *n* with replacement. If the two parental strains are the same, the original strain is transmitted. If different, they recombine, and the recombinant strain is transmitted. The probabilities of transmitting the original or recombinant strain are 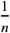 and 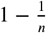, respectively.

Given that orthologous gene pairs between two parental strains are often unknown, we implement meiotic recombination by randomly selecting genes from the pooled genes of the two strains. This assumption is reasonable as physical locations of *var* genes can be mobile through ectopic recombination and gene conversions. The resulting offspring strains share some fraction of their *var* genes with the parental strains.

#### Within-host dynamics

Each strain is individually tracked through its entire life cycle, encompassing the liver stage and asexual blood stage in the human host, and the sexual stage in the mosquito. As we do not explicitly model mosquitoes, we delay the expression of each strain in the recipient host by 7 days to account for the sexual stage, e.g., the time required for gametocytes to develop into sporozoites within mosquitoes. Additionally, we delay the expression of each strain by another 7 days to account for the liver stage, e.g., the time required for parasites to be released as merozoites into the bloodstream to invade red blood cells. Hence, after a total of 14 days, the active asexual bloodstage and the expression of the *var* repertoire begin.

The *var* genes within an infection are expressed sequentially (***Zhang and Deitsch, 2022***; ***Deitsch and Dzikowski, 2017***). During this process, the host is considered infectious with the currently active strain, and only these blood-stage strains are transmissible to another host. The deactivation rate of each gene is governed by the host’s variant-specific immunity. When a gene is actively expressed, the host’s immune system “checks” whether either of its two epitopes has been encountered previously-either during past infections or earlier in the current infection through genes that have already been expressed. If both epitopes are novel to the host, it takes approximately seven days for the immune system to mount an effective antibody response, after which the expressed variant rapidly declines and is cleared(***Gatton and Cheng, 2004***). Prior exposure to one epitope shortens the expression duration by about half, whereas prior exposure to both epitopes results in immediate deactivation. Thus, the active period of a gene is proportional to the number of its epitopes unseen by the host. Once a gene is deactivated, its two epitopes are added to the host’s immune memory, and the next new gene in the repertoire becomes active. The strain is cleared once its entire *var* gene repertoire has been expressed or recognized. Consequently, the total infection duration of a given repertoire is proportional to the total number of previously unseen epitopes across its *var* genes. Immunity to a given epitope wanes over time (***COLLINS et al., 1968***; ***Collins et al., 1964***), requiring re-exposure for maintenance.

We also consider a variation on the within-host rules in addition to the baseline assumption that an infection clears only after the parasite has exhausted its entire *var* gene repertoire. This modification is motivated by biological evidence that clearance can occur earlier for several reasons, including stochastic extinction before full repertoire exhaustion. Even if some *var* genes remain unexpressed, an infection may terminate once parasite densities fall to very low levels due to demographic stochasticity. Such declines in parasite densities can arise from non-variant-specific immune mechanisms or from cross-immunity among *var* genes that share sequence similarity or epitopes (***Holding and Recker, 2015***; ***Crompton et al., 2014***; ***Langhorne et al., 2008b***), both of which can substantially reduce parasite numbers. Here, we focus on the latter. To capture the possibility of early termination, we implemented a simple scenario in which there is a small probability of clearing the current infection while any given *var* gene-whether non-final or final-is being expressed. This probability is determined by the host’s pre-existing immunity to the two epitopes (alleles) of that gene, thereby capturing in a parsimonious manner the effects of cross-immunity among sequence- or allele-sharing *var* genes in reducing parasitemia. Specifically, it is modeled as a Bernoulli draw whose success probability equals the immunity level against the gene (0 for no immunity to either epitope, 0.5 for immunity to one epitope, and 1 for immunity to both epitopes) multiplied by a constant factor of 0.025. Thus, the probability scales with pre-exisiting variant-specific immunity to the gene but remains small overall, while introducing additional variance into the emergent distribution of total infection duration across hosts.

We note that in the ABM simulations, we do not use empirical estimates of infection duration from the historical neurosyphilis treatment studies of immunologically naïve individuals as direct inputs. Instead, infection duration emerges from the within-host dynamics described in the previous paragraph. In our simulations, we assume deactivation times of 7, 7.5, or 8 days for a gene to which the host is naive. These values are consistent with the duration of each successive parasitemia peak observed in *Plasmodium falciparum* infections, each peak corresponding primarily to the expression of a single *var* gene. These mean expression durations, in combination with the within-host rules described previously, produce distributions of infection duration across naïve hosts and those aged 1-5 years that span means and variances above and below, but collectively encompassing, values comparable to the historical clinical data from naïve neurosyphilis patients treated with *P. falciparum* malaria. We provide a set of illustrative supplementary figures showing that the simulated distributions of infection duration overlap with and closely resemble the empirical distribution from the historical clinical data (Appendix 1-Figure 27-32).

We acknowledge that the ABM cannot capture all mechanisms and complexities of within-host malaria dynamics, many of which remain poorly understood. Nonetheless, the model generates a range of distributions of infection duration spanning distributions that closely match those from the historical clinical data. Because the queueing-theory methods rely on the mean and variance of infection duration to infer FOI, this range of scenarios provides an appropriate basis for evaluating their performance.

We consider carrying capacities for both liver- and blood-stage infection (***Tiedje et al., 2025***), adopting a value of 30 for both based on reasons detailed in Materials and Methods-Choice of *c* and *r* for the *GI*/*G*/*c*/*c* + *r* queue in the two-moment approximation method. When the number of liver-stage strains reaches the specified carrying capacity, the host no longer receives additional infections if selected as the recipient host for transmission events. Similarly, when the number of blood-stage strains reaches the carrying capacity, liver-stage strains are not released into the bloodstream and fail to transition to the blood-stage, effectively being lost.

Details of the parameters and their values are summarized in the supplementary file 5-simParams.xlsx.

### Experimental designs

Each simulation runs for either 200 years (closed systems) or 150 years (open systems) to reach a semi-stationary state before introducing transmission-reducing interventions. We simulate malaria dynamics with parameters representative of high-transmission endemic regions, considering both constant and seasonal transmission, and across different spatial configurations, including closed, semi-open, and regionally-open systems.

We consider indoor residual spraying (IRS), which involves applying insecticide to the internal walls and ceilings of homes (***World Health Organization, 2015***) in the field. IRS effectively reduces the mosquito population, thereby decreasing the transmission rate. We simulate three temporary IRS interventions with varying coverages: low (reducing transmission by around 20%), mid (reducing transmission by around 40-45%), and high (reducing transmission by around 65-75%). Each IRS intervention lasts for three years.

Seasonality is represented as a scaling constant multiplied by a temporal vector of 360 days, which represents the daily number of mosquitoes over a year. This temporal vector (***Pilosof et al., 2019***) is derived from a deterministic model of mosquito population dynamics (***White et al., 2011***). The model, originally developed for *Anopheles gambiae*, includes a set of ordinary differential equations describing the dynamics of four mosquito stages: eggs, larvae, pupae, and adults. Seasonality is implemented via density dependence at the egg and larva stages as a function of rainfall (availability of breeding sites). For our purposes, the values of the effective contact rate are more critical than the absolute number of daily mosquitoes. Essentially, we have a basic vector that represents the number of mosquitoes throughout the year and a scaling constant that encapsulates all other parameters related to vectorial capacity. The product of this temporal vector and the scaling constant results in the effective contact rate.

In closed systems, migration is discontinued after the initial seeding of local transmission from a regional pool of *var* genes. In semi-open systems, two individual populations are explicitly coupled via migration. Regionally-open systems involve a local population with migration from a regional pool, which acts as a proxy for regional parasite diversity, i.e., diversity from the aggregated individual populations in the region. Because each parasite genome is a repertoire with a given number of *var* genes, migrant genomes are assembled by randomly sampling *var* genes from the regional pool.

Transmission can be homogeneous or heterogeneous across individual hosts. For heterogeneity, we consider two groups of human hosts: a high risk group, which receives approximately 94% of the bites, and a low risk group, which receives the remaining fraction. The high-risk group is twice as large as the low-risk group (2:1 ratio).

## Test of deviation from Poisson homogeneity in MOI estimates

Many studies have explored whether a count dataset deviates significantly from a homogeneous Poisson distribution. We use the Potthoff-Whittinghill “index of dispersion” test (***Potthoff and Whittinghill, 1966***; ***Lloyd-Smith et al., 2005***), which is asymptomatically locally most powerful against the negative binomial alternative. For a dataset with elements, the statistic is 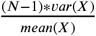 and its asymptotic distribution is chi-squared with *N* −1 degrees of freedom. The p-value is the cumulative density of the chi-squared (*N* − 1) distribution to the right of the test statistic, representing the probability that the observed variance could have arisen by chance from a Poisson distribution.

We summarize the p-values of the Potthoff-Whittinghill “index of dispersion” test in supplementary file 2-deviationFromPoissonTest.xlsx. The p-values are close to 0, indicating a significant deviation from a Poisson distribution for both simulated outputs and empirical surveys in Bongo District. Few exceptions occur in high-coverage IRS scenarios in the simulated outputs, where prevalence is very low and most infections are mono-clonal.

## Reducing stochastic impact in sampling processes

To mitigate the effects of stochasticity in various sampling processes within the analysis, we perform 200 realizations, including the incorporation of the measurement error model into the simulation output. Each realization results in a slightly different collection of subsampled *var* genes per host, potentially leading to minor variations in individual MOI estimates. To impute MOI estimates for treated individuals or those with missing data, we conduct 200 samplings from available nontreated individuals. We then calculate a final population-level MOI distribution, weighted across these 200 realizations or samplings. This approach reduces the impact of extreme single-sampling variations on MOI estimates and FOI inference.

## Confidence intervals for FOI inference

### Non-parametric bootstrap

Bootstrap datasets are generated by resampling with replacement from the original population-level MOI distribution, i.e., the collection of individual MOI estimates, using individual-level MOI as the unit of the bootstrap sampling. We run 200 bootstrap replicates, as this number has been tested and shown to achieve a coefficient of variation comparable to that obtained with a higher number of replicates (***Efron and Tibshirani, 1994***). For each bootstrap dataset, we determine the maximum likelihood estimates for FOI. The 200 replicates thus produce a bootstrap sampling distribution for FOI estimates. We calculate the skewness of these distributions. The majority fall within the range of -0.5 to 0.5, with a few exceptions falling within the range of 0.5-0.75. Therefore, we consider them fairly symmetric and do not apply a skewness adjustment to ensure good coverage. Detailed skewness values can be found in supplementary file 6-FOIBootstrapSkewness.xlsx.

## A comparison of the performance of the Bayesian formulation and the original *var*coding method based on the simulation output

We compare the performance of the original *var*coding method and its Bayesian formulation against true MOI values using the Cramer-von Mises and Anderson-Darling tests.

MOI estimates from the *var*coding method often differ significantly from true MOI values (p-value < 0.05). Similarly, there is frequently a statistically significant difference between MOI estimates from the *var*coding method and those from the Bayesian formulation. However, differences between MOI estimates from the Bayesian formulation and true MOI values are generally not statistically significant.

Therefore, the Bayesian formulation improves upon the original *var*coding method, which assumes a constant repertoire size to estimate MOI. This improvement is expected, as the Bayesian formulation accounts for variation in repertoire size due to *var* gene under-sampling.

Both methods perform well in low-transmission settings, where true MOI values are low. In moderate and high-transmission scenarios, the *var*coding method remains reasonably effective, but the Bayesian formulation demonstrates a notable improvement in capturing higher MOI values, where measurement error is more pronounced. This improvement is significant in the high-transmission endemic Bongo District of Ghana, where our empirical MOI estimates were derived.

The documented test results are included in supplementary file 7-BayesianImprovement.xlsx. Overall, both methods tend to underestimate MOI. While the Bayesian formulation provides a more accurate and robust estimation by addressing the imperfect detection of *var* genes, neither method accounts for other factors that may reduce the number of *var* genes detected per individual (see Discussion).

**Figure 11.**
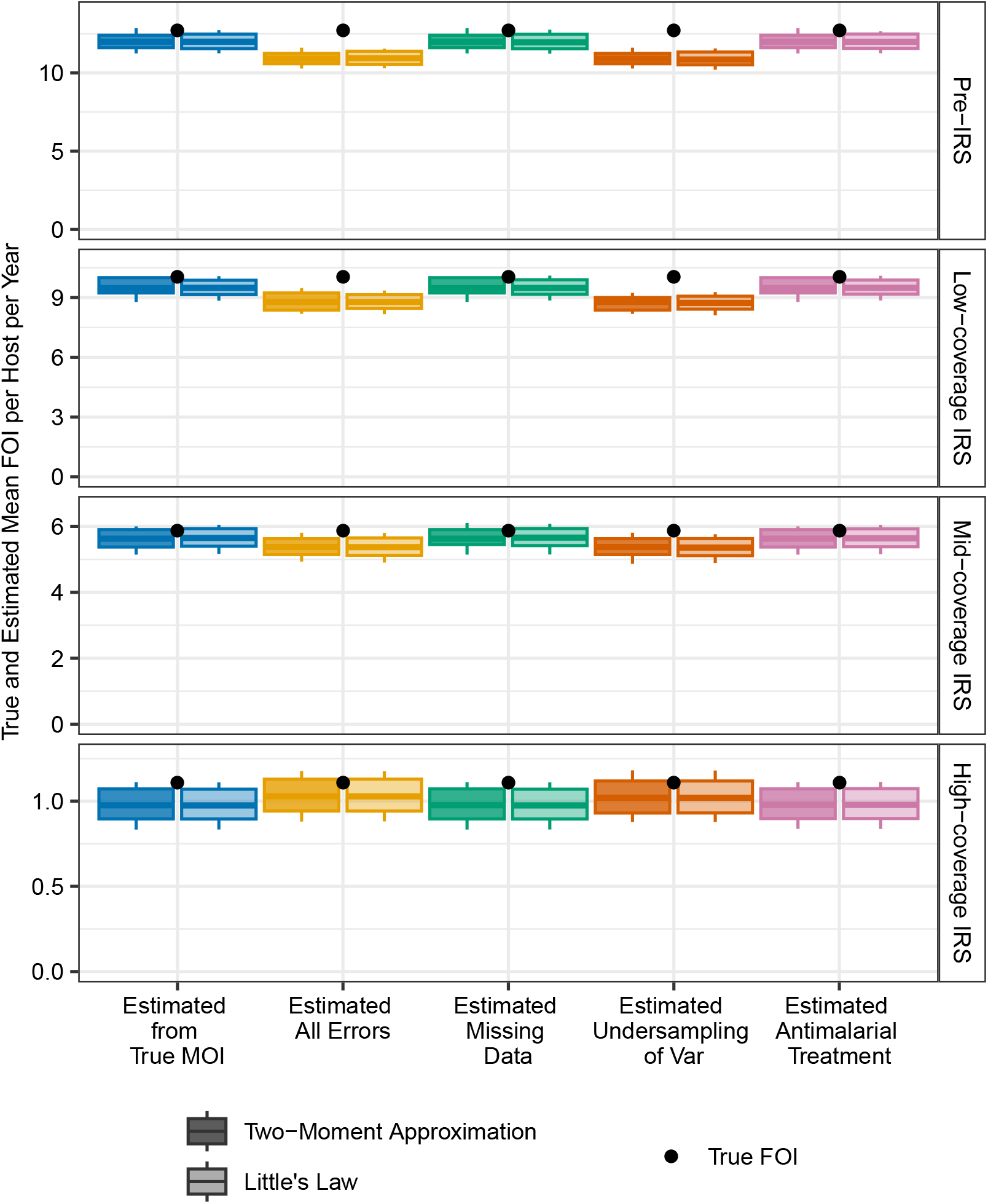
True and estimated FOI by the two-moment and Little’s Law methods for additional simulated scenarios of homogeneous exposure risk. The times between local transmission events are Gamma-distributed, with non-seasonal transmission in a closed system. The true mean FOI per host per year is calculated by dividing the total number of infections acquired by the population by the total number of hosts in the population. Each boxplot shows minimum, 5% quantile, median, 95% quantile, and maximum values.

**Figure 12.**
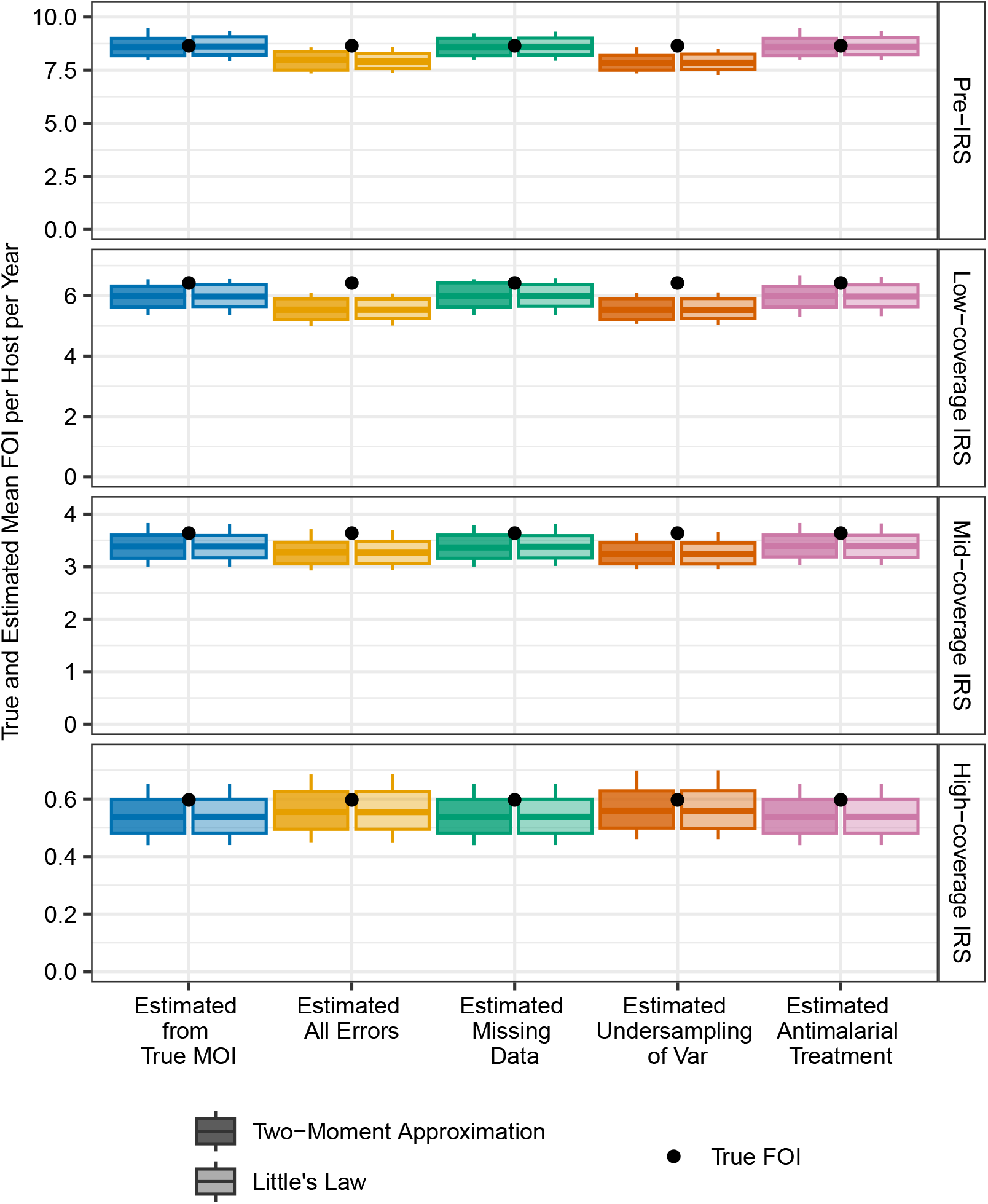
True and estimated FOI by the two-moment and Little’s Law methods for additional simulated scenarios of heterogeneous exposure risk. The times between local transmission events are Gamma-distributed, with non-seasonal transmission in a semi-open system. The true mean FOI per host per year is calculated by dividing the total number of infections acquired by the population by the total number of hosts in the population. Each boxplot shows minimum, 5% quantile, median, 95% quantile, and maximum values.

**Figure 13.**
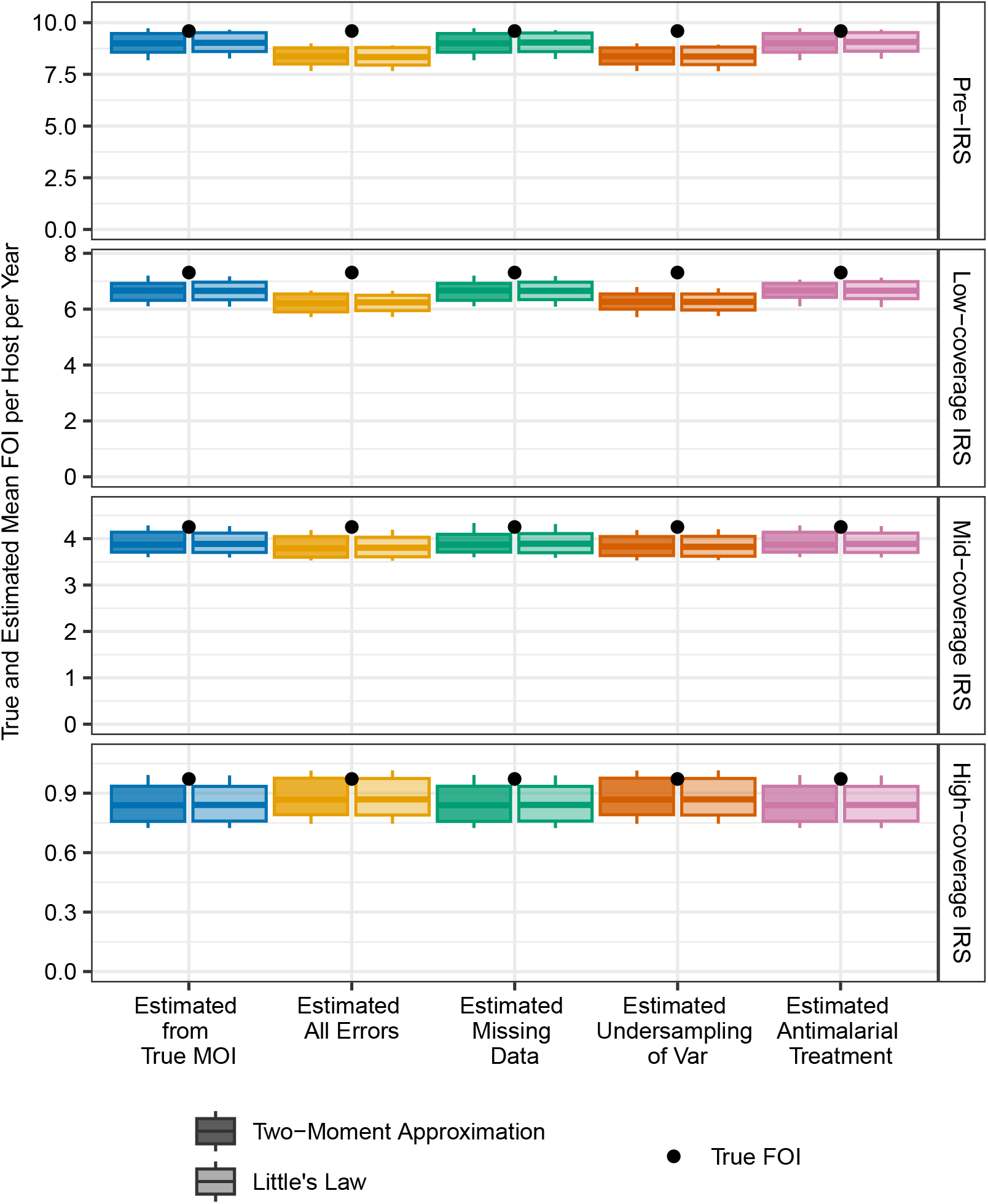
True and estimated FOI by the two-moment and Little’s Law methods for additional simulated scenarios of homogeneous exposure risk. The times between local transmission events are Gamma-distributed, with seasonal transmission in a regionally-open system. The true mean FOI per host per year is calculated by dividing the total number of infections acquired by the population by the total number of hosts in the population. Each boxplot shows minimum, 5% quantile, median, 95% quantile, and maximum values.

**Figure 14.**
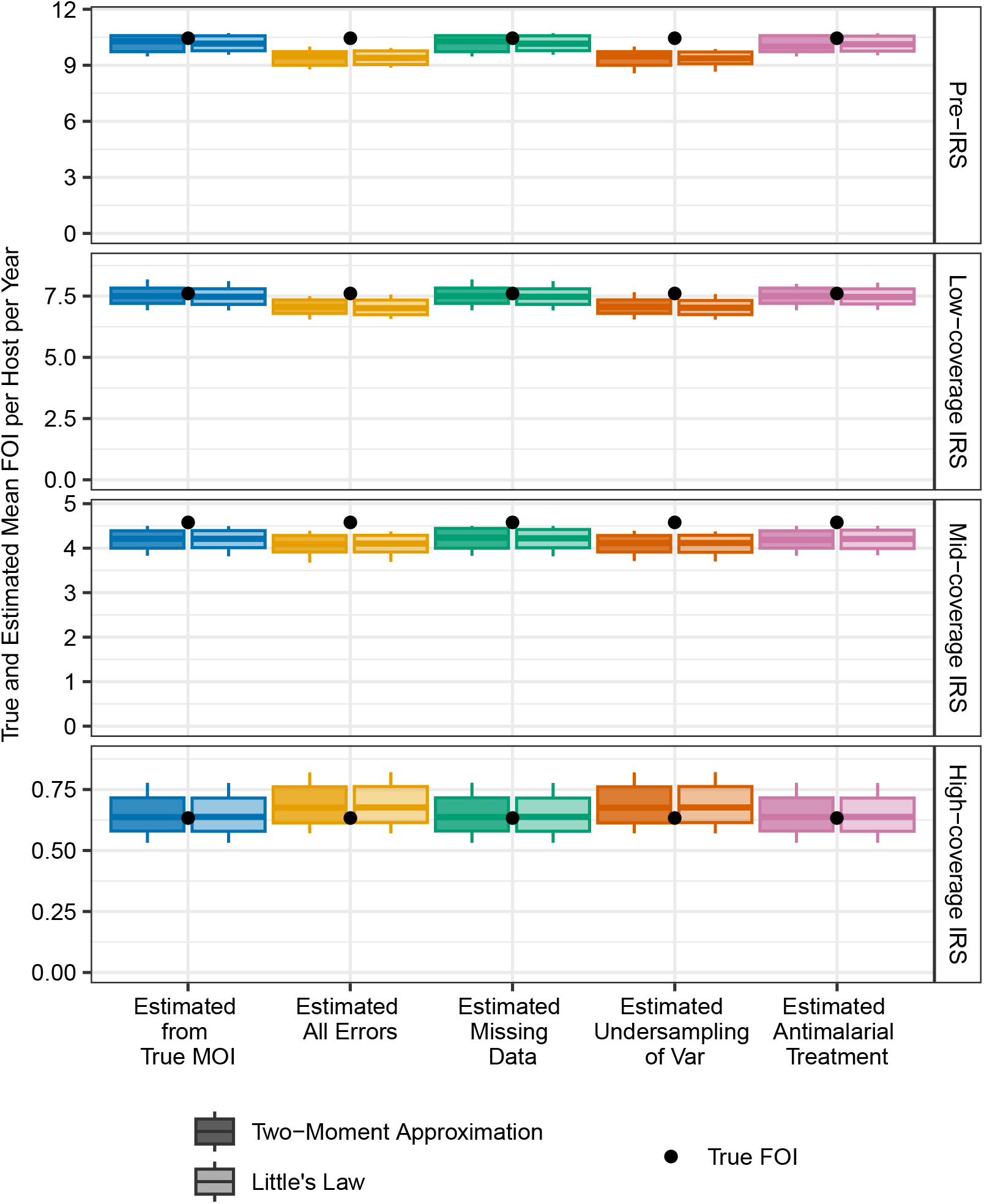
True and estimated FOI by the two-moment and Little’s Law for additional simulated scenarios of homogeneous exposure risk. The times between local transmission events are Gamma-distributed, with non-seasonal transmission in a regionally-open system. The true mean FOI per host per year is calculated by dividing the total number of infections acquired by the population by the total number of hosts in the population. Each boxplot shows minimum, 5% quantile, median, 95% quantile, and maximum values.

**Figure 15.**
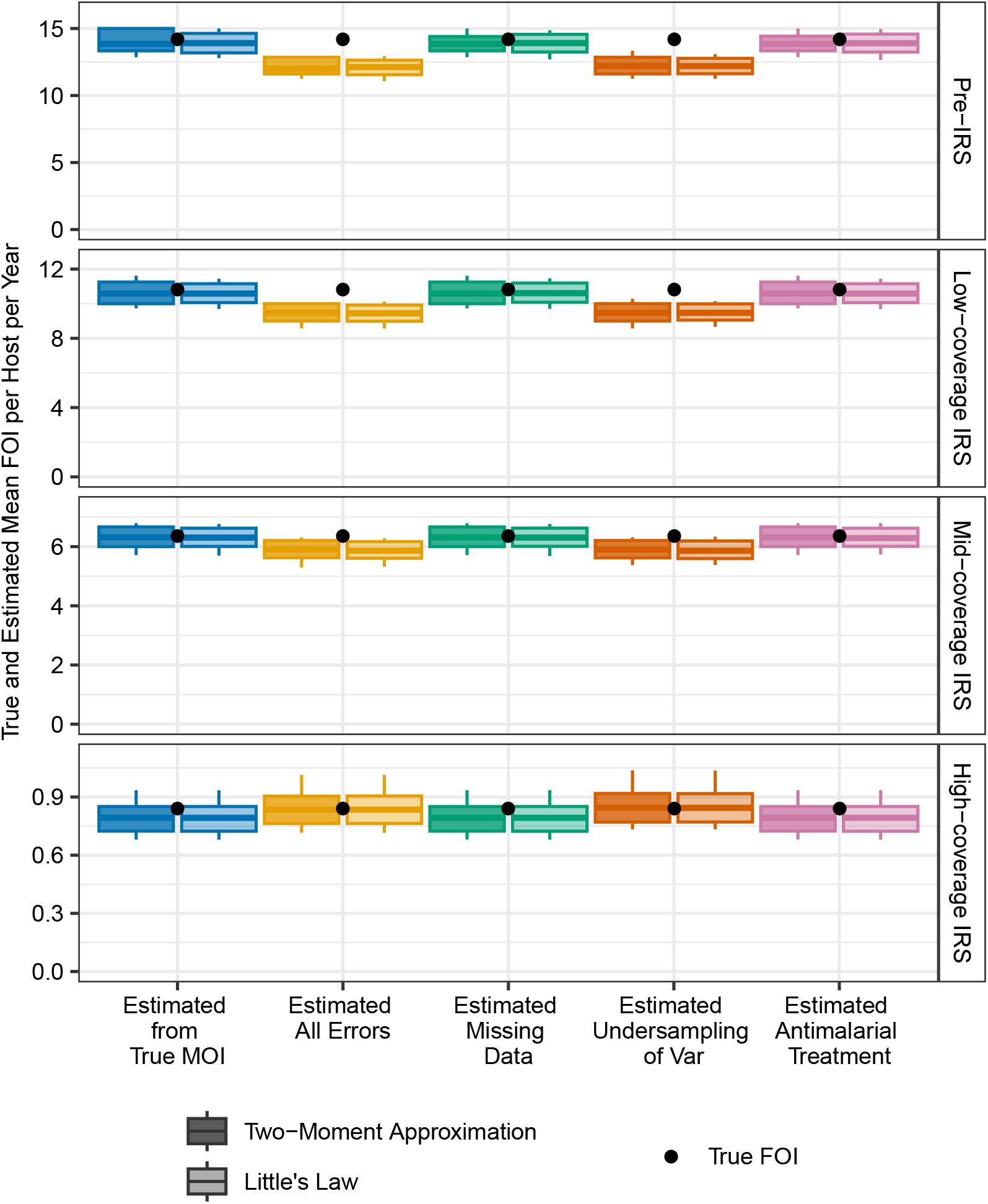
True and estimated FOI by the two-moment and Little’s Law methods for additional simulated scenarios of homogeneous exposure risk. The times between local transmission events follow a exponential distribution, with seasonal transmission in a closed system. The true mean FOI per host per year is calculated by dividing the total number of infections acquired by the population by the total number of hosts in the population. Each boxplot shows minimum, 5% quantile, median, 95% quantile, and maximum values.

**Figure 16.**
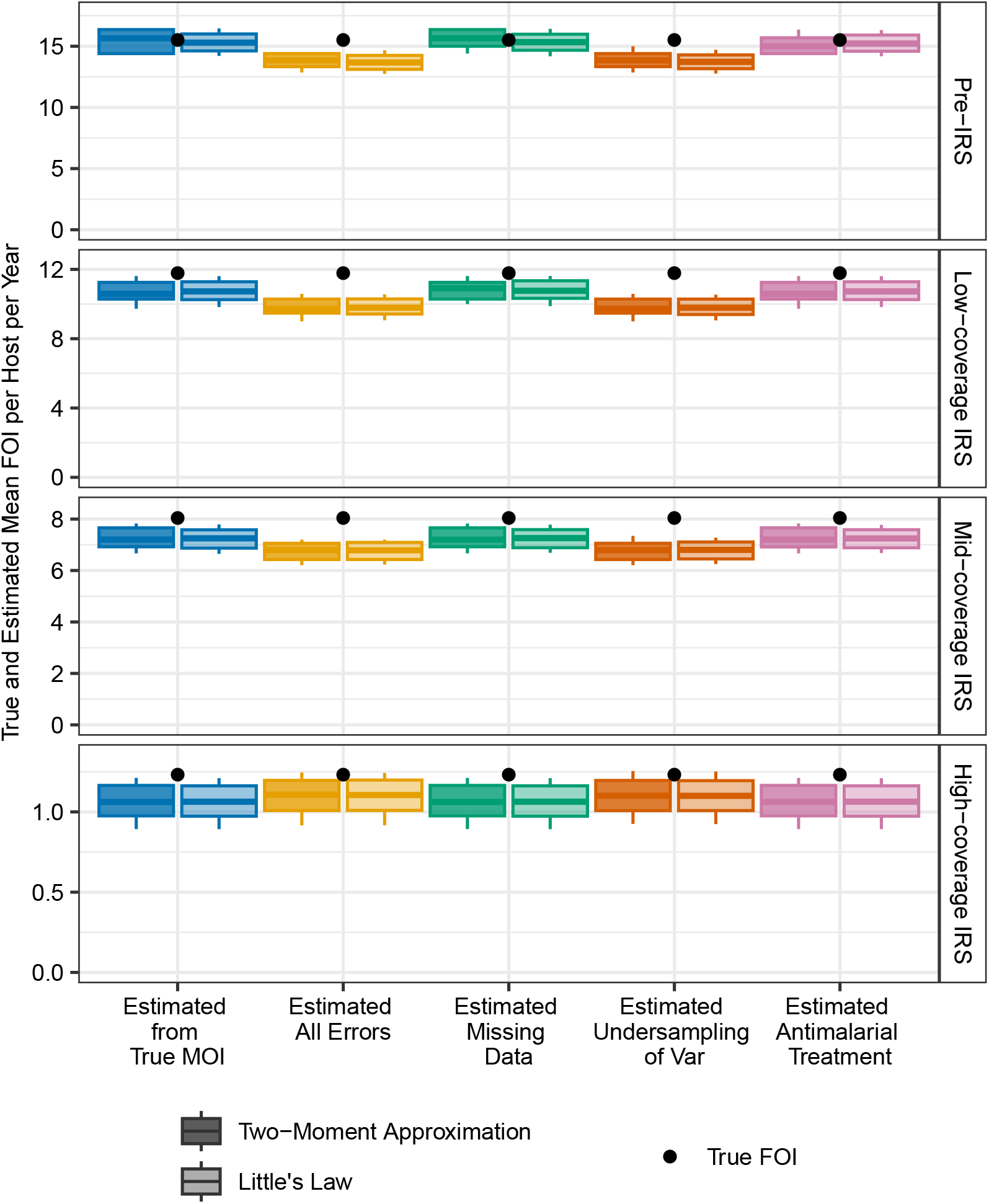
As in Figure 1, we present confidence intervals for the estimated mean FOI values; all aspects of the simulation setup are identical except that infections are allowed to clear stochastically before full repertoire exhaustion. Specifically, while any *var* gene, whether non-final or final, is being expressed, there is a small probability of infection clearance that depends on the host’s pre-existing immunity to that gene’s epitopes (Appendix 1–Simulation data, subsection “An extended *var* model,” sub-subsection “Within-host dynamics”).

**Figure 17.**
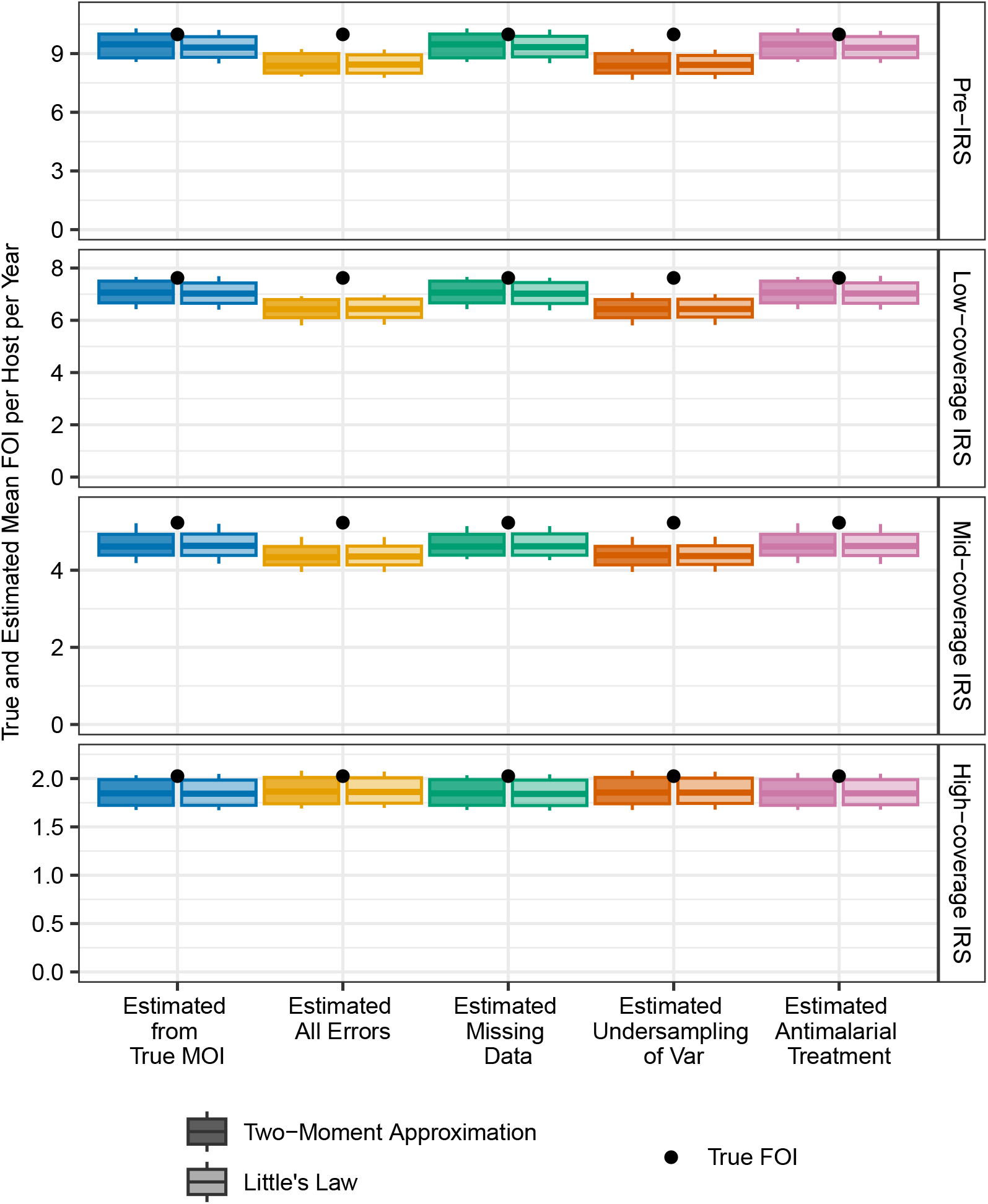
As in Figure 2, we present confidence intervals for the estimated mean FOI values; all aspects of the simulation setup are identical except that infections are allowed to clear stochastically before full repertoire exhaustion. Specifically, while any *var* gene, whether non-final or final, is being expressed, there is a small probability of infection clearance that depends on the host’s pre-existing immunity to that gene’s epitopes (Appendix 1–Simulation data, subsection “An extended *var* model,” sub-subsection “Within-host dynamics”).

**Figure 18.**
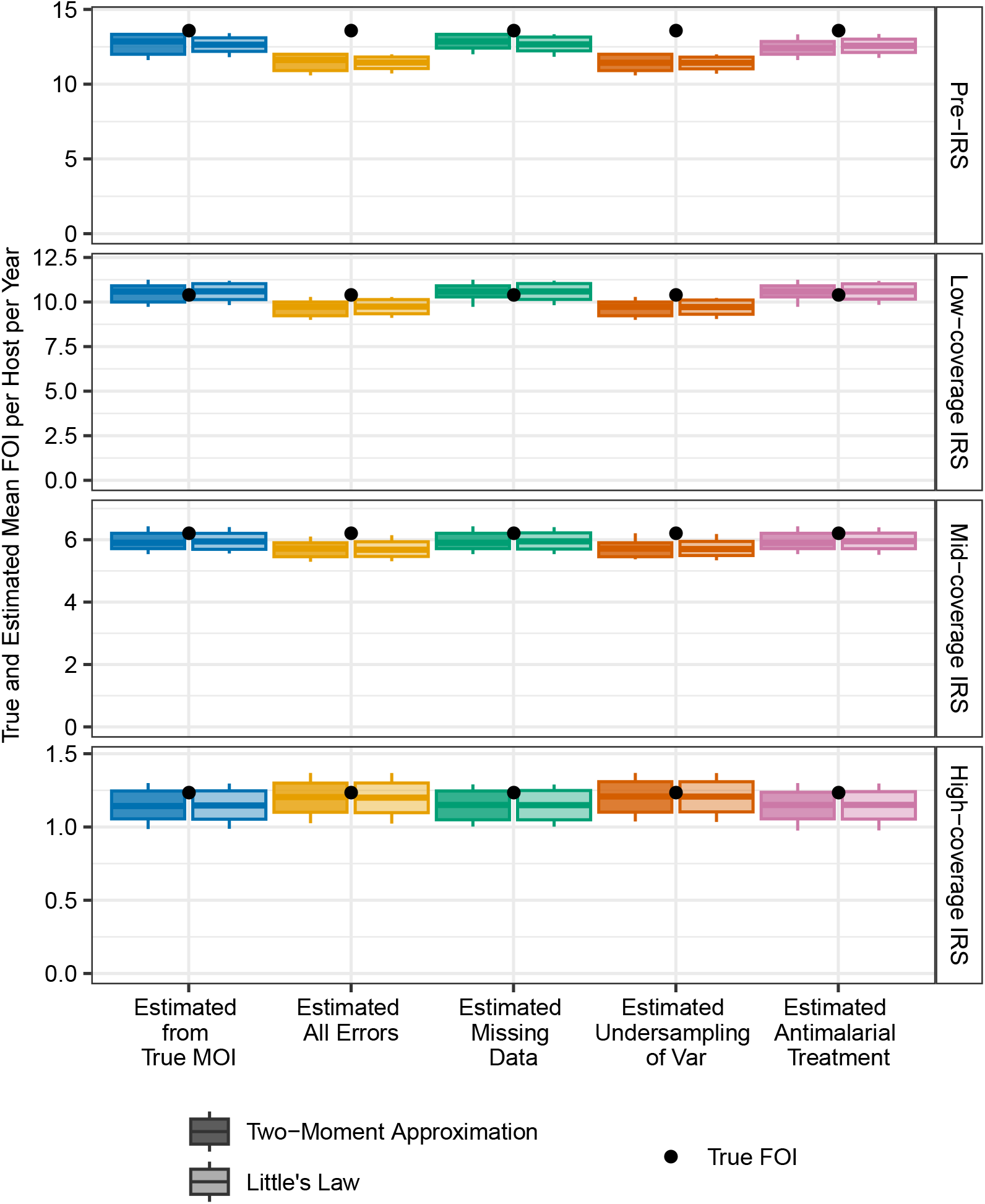
As in Appendix 1-Figure 11, we present confidence intervals for the estimated mean FOI values; all aspects of the simulation setup are identical except that infections are allowed to clear stochastically before full repertoire exhaustion. Specifically, while any *var* gene, whether non-final or final, is being expressed, there is a small probability of infection clearance that depends on the host’s pre-existing immunity to that gene’s epitopes (Appendix 1–Simulation data, subsection “An extended *var* model,” sub-subsection “Within-host dynamics”).

**Figure 19.**
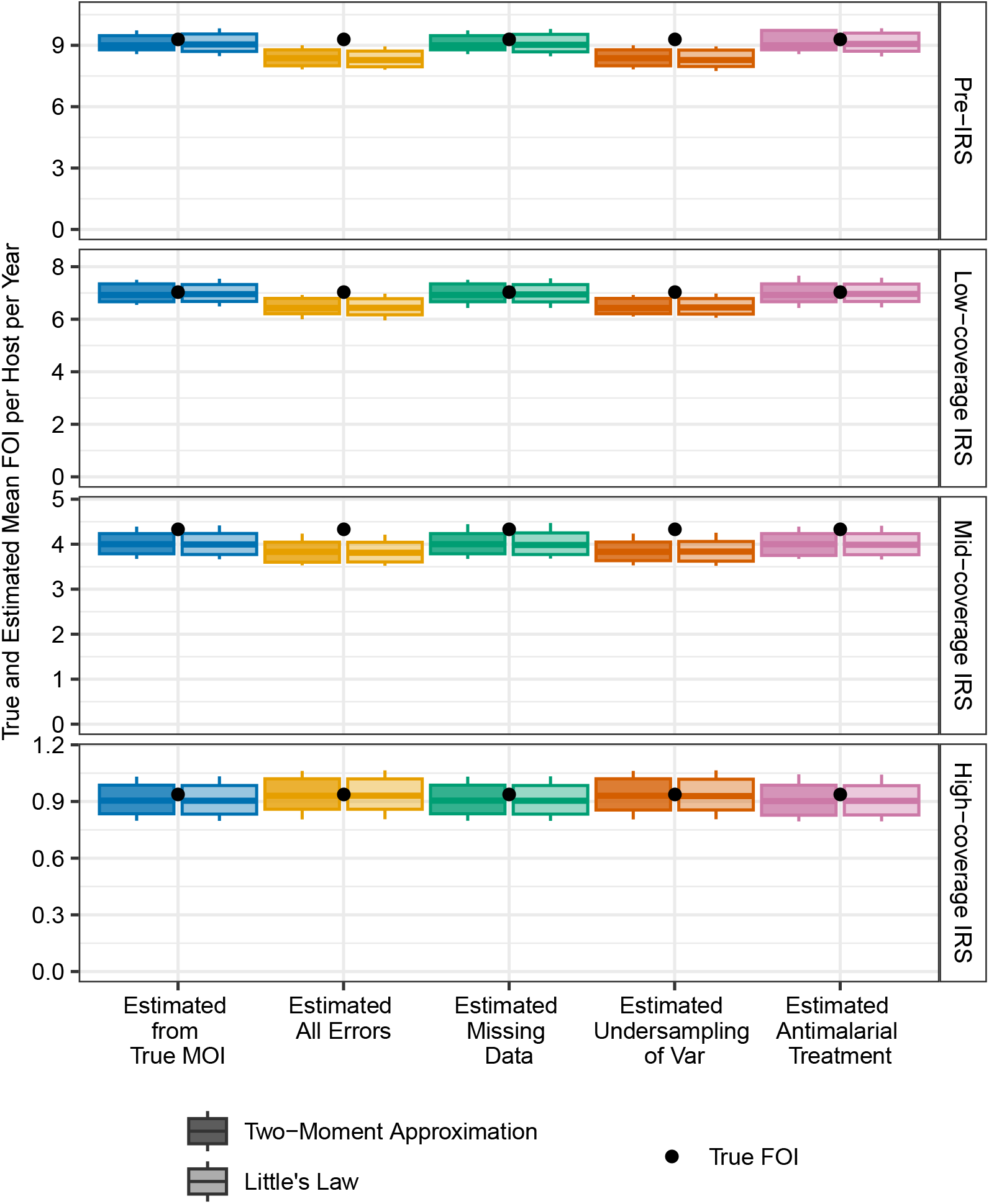
As in Appendix 1-Figure 12, we present confidence intervals for the estimated mean FOI values; all aspects of the simulation setup are identical except that infections are allowed to clear stochastically before full repertoire exhaustion. Specifically, while any *var* gene, whether non-final or final, is being expressed, there is a small probability of infection clearance that depends on the host’s pre-existing immunity to that gene’s epitopes (Appendix 1–Simulation data, subsection “An extended *var* model,” sub-subsection “Within-host dynamics”).

**Figure 20.**
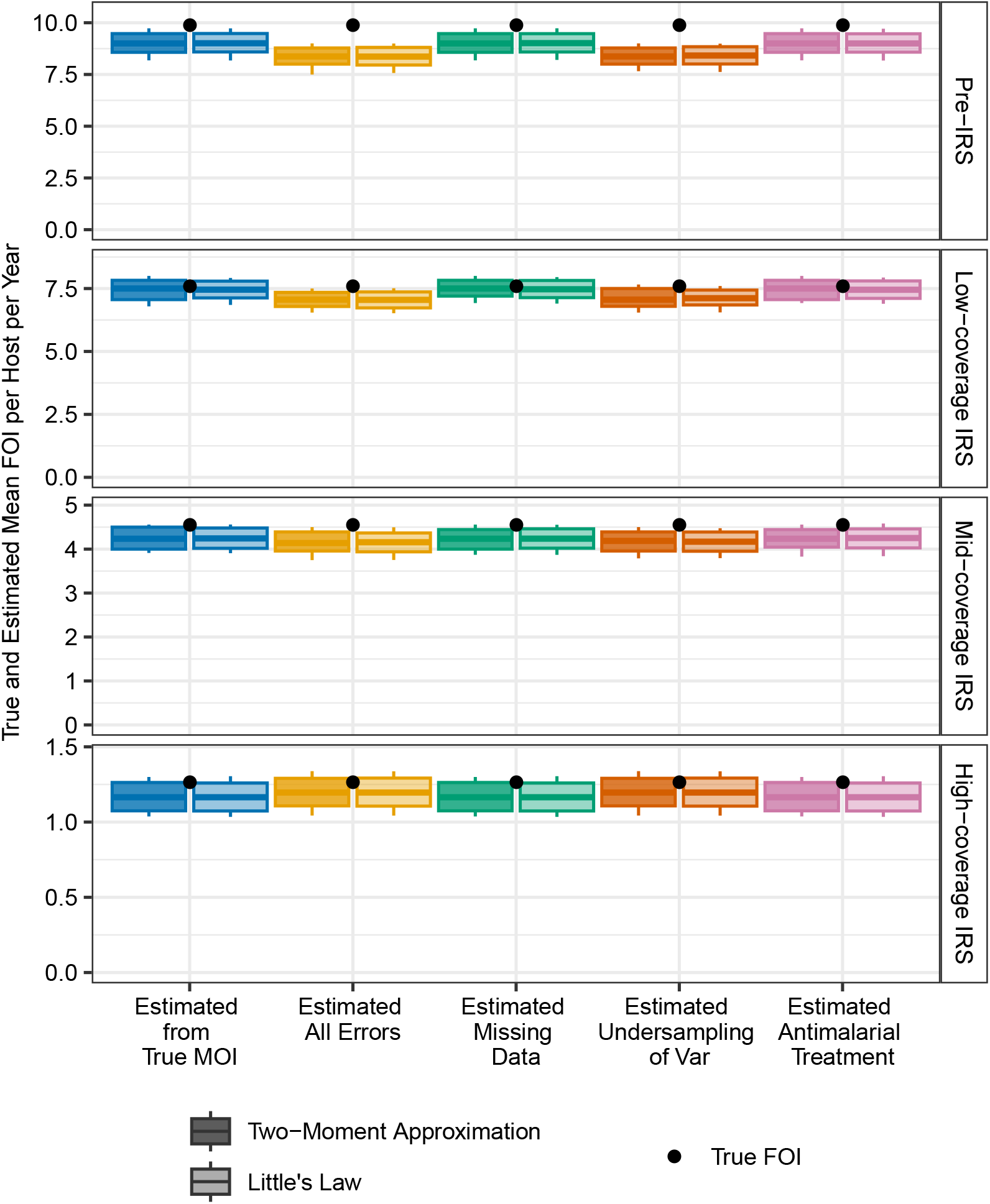
As in Appendix 1-Figure 13, we present confidence intervals for the estimated mean FOI values; all aspects of the simulation setup are identical except that infections are allowed to clear stochastically before full repertoire exhaustion. Specifically, while any *var* gene, whether non-final or final, is being expressed, there is a small probability of infection clearance that depends on the host’s pre-existing immunity to that gene’s epitopes (Appendix 1–Simulation data, subsection “An extended *var* model,” sub-subsection “Within-host dynamics”).

**Figure 21.**
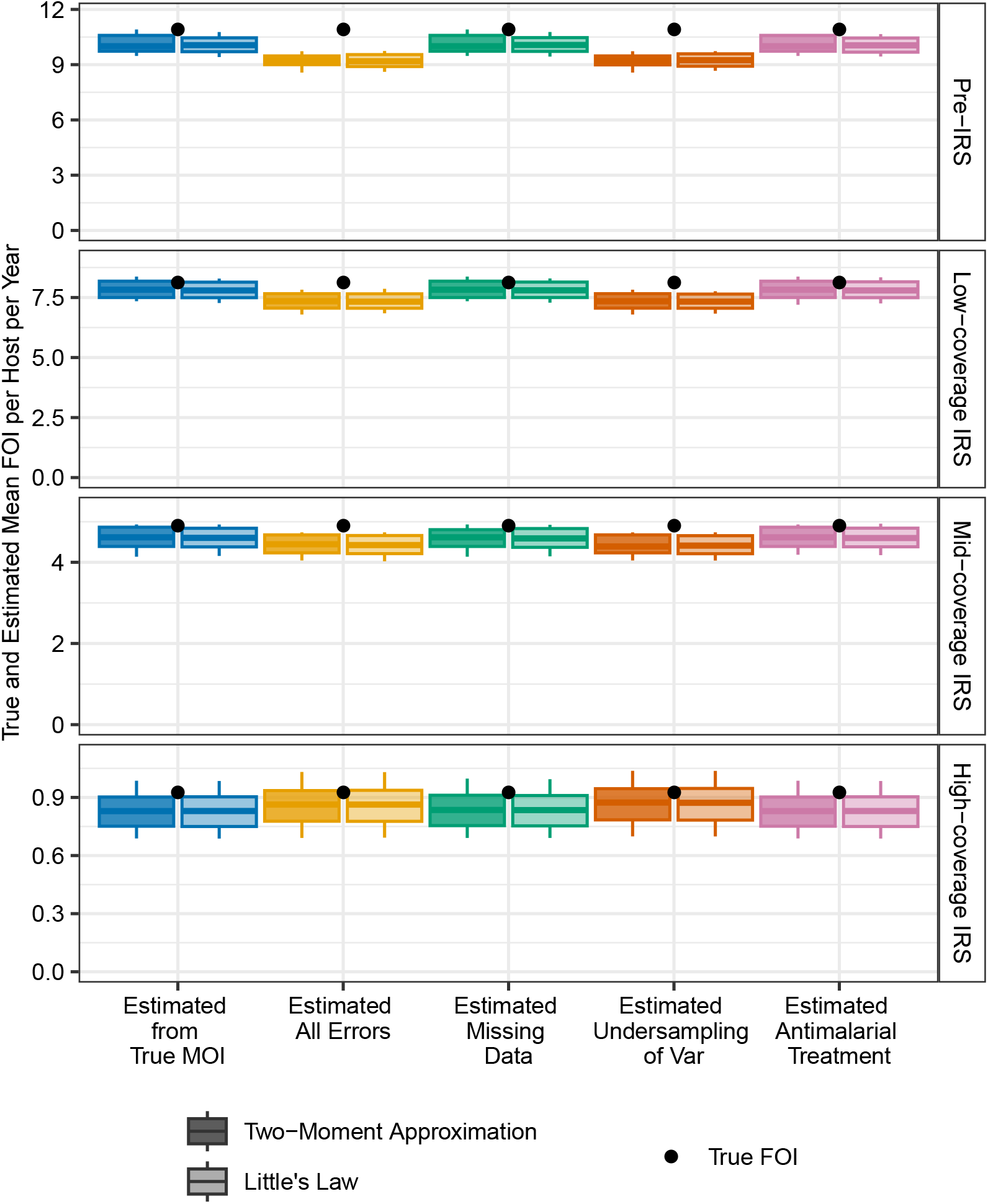
As in Appendix 1-Figure 14, we present confidence intervals for the estimated mean FOI values; all aspects of the simulation setup are identical except that infections are allowed to clear stochastically before full repertoire exhaustion. Specifically, while any *var* gene, whether non-final or final, is being expressed, there is a small probability of infection clearance that depends on the host’s pre-existing immunity to that gene’s epitopes (Appendix 1–Simulation data, subsection “An extended *var* model,” sub-subsection “Within-host dynamics”).

**Figure 22.**
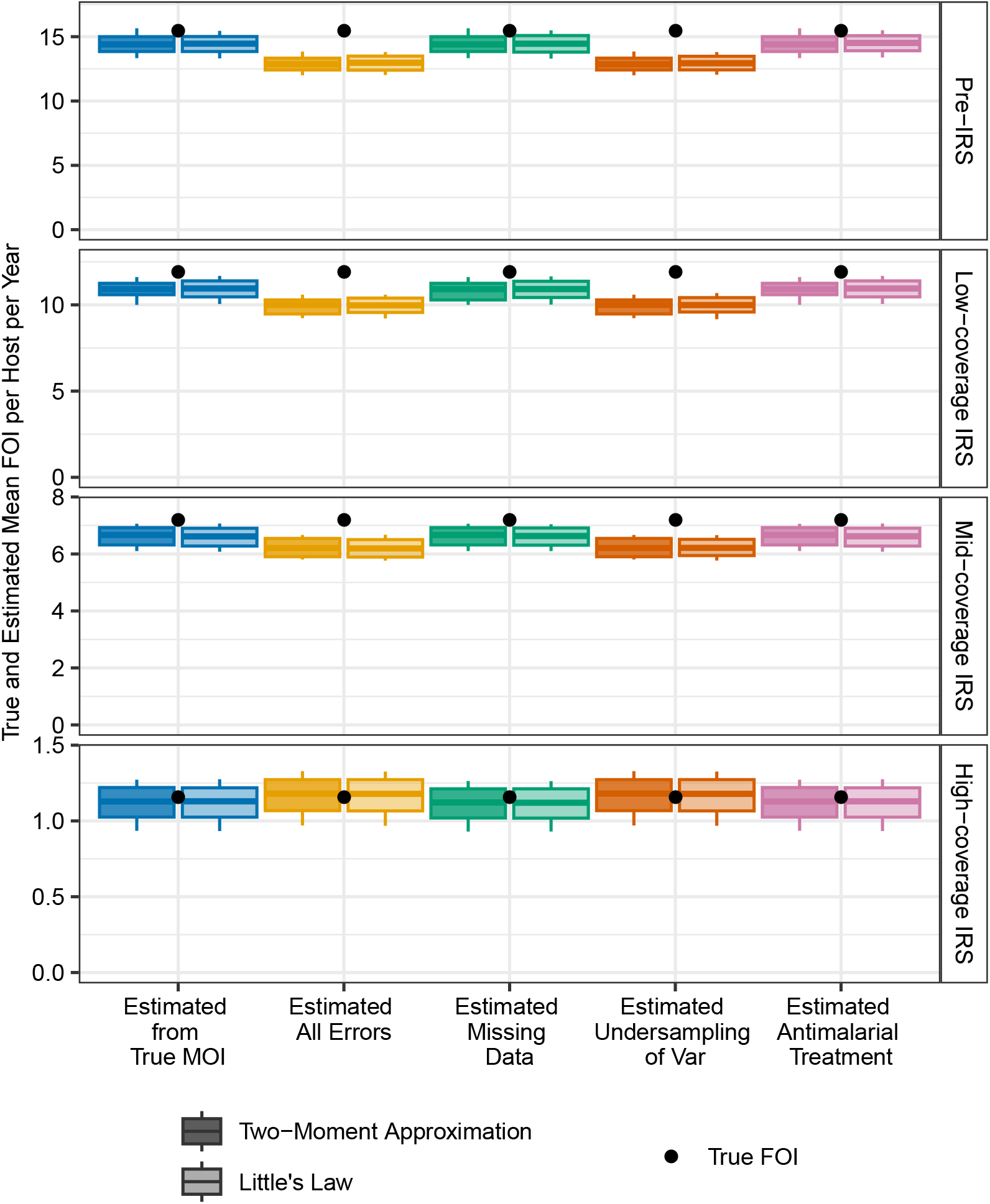
As in Appendix 1-Figure 15, we present confidence intervals for the estimated mean FOI values; all aspects of the simulation setup are identical except that infections are allowed to clear stochastically before full repertoire exhaustion. Specifically, while any *var* gene, whether non-final or final, is being expressed, there is a small probability of infection clearance that depends on the host’s pre-existing immunity to that gene’s epitopes (Appendix 1–Simulation data, subsection “An extended *var* model,” sub-subsection “Within-host dynamics”).

**Figure 23.**
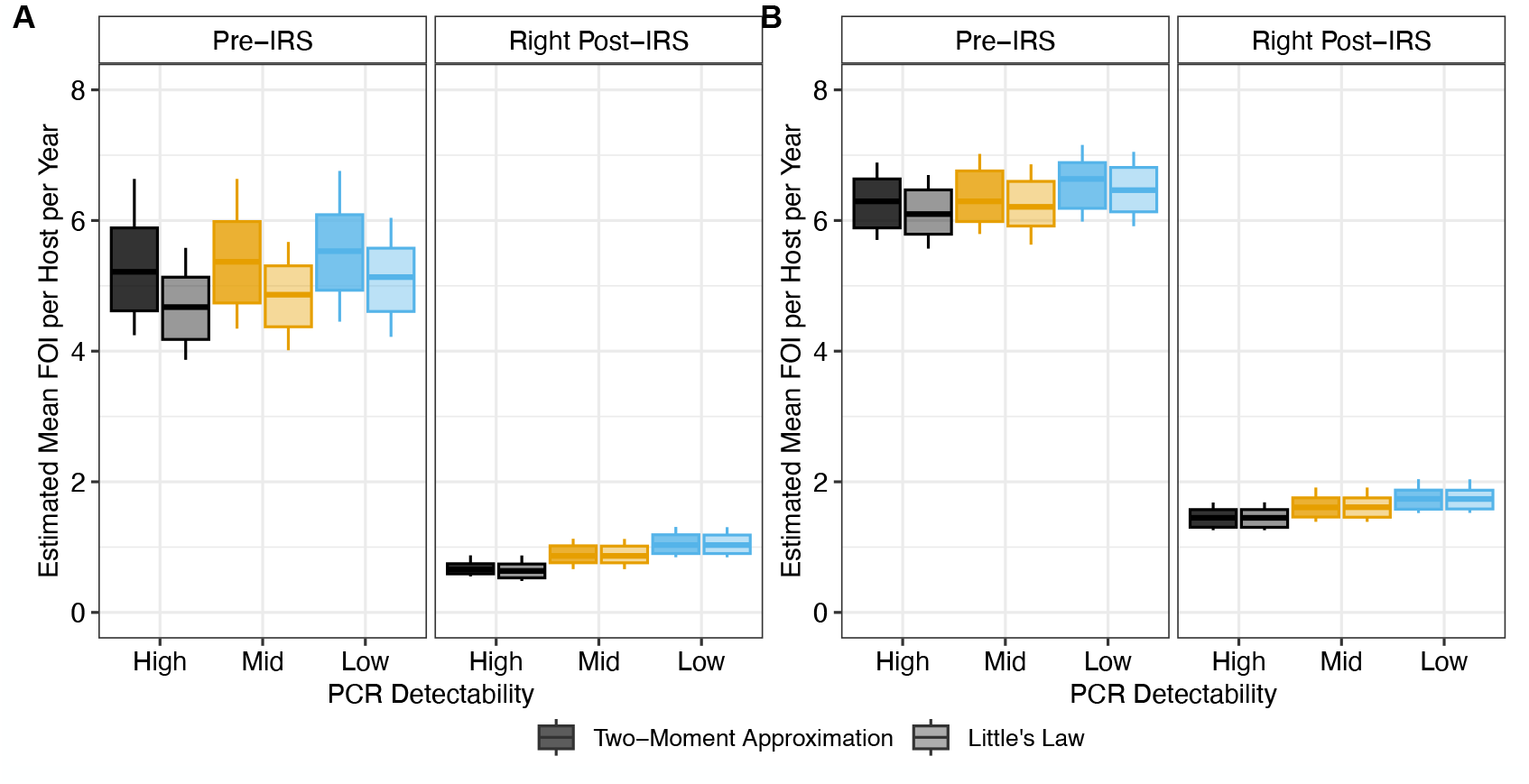
Confidence intervals for the estimated mean FOI values in Ghana surveys before and immediately after a transient three-round IRS intervention (A) The estimated FOI values when excluding these treated individuals from the analysis. (B) The estimated FOI values when discarding the infection status and MOI estimates of treated individuals and sampling from non-treated ones. Each boxplot shows minimum, 5% quantile, median, 95% quantile, and maximum. The value of *c* is set to 25.

**Figure 24.**
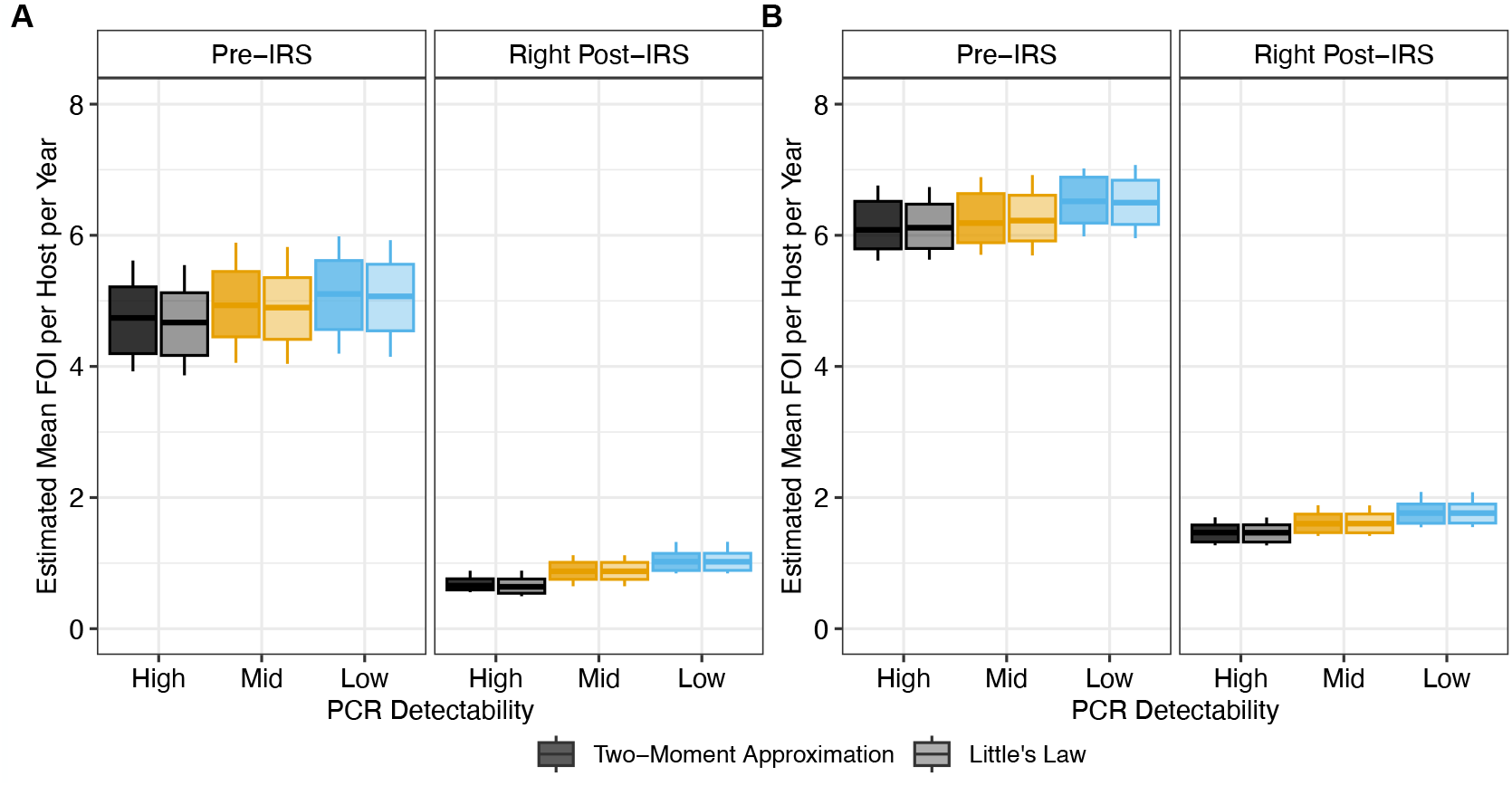
Confidence intervals for the estimated mean FOI values in Ghana surveys before and immediately after a transient three-round IRS intervention (A) The estimated FOI values when excluding these treated individuals from the analysis altogether. (B) The estimated FOI values when discarding the infection status and MOI estimates of treated individuals and sampling from non-treated ones. Each boxplot shows minimum, 5% quantile, median, 95% quantile, and maximum. The value of *c* is set to 40.

**Figure 25.**
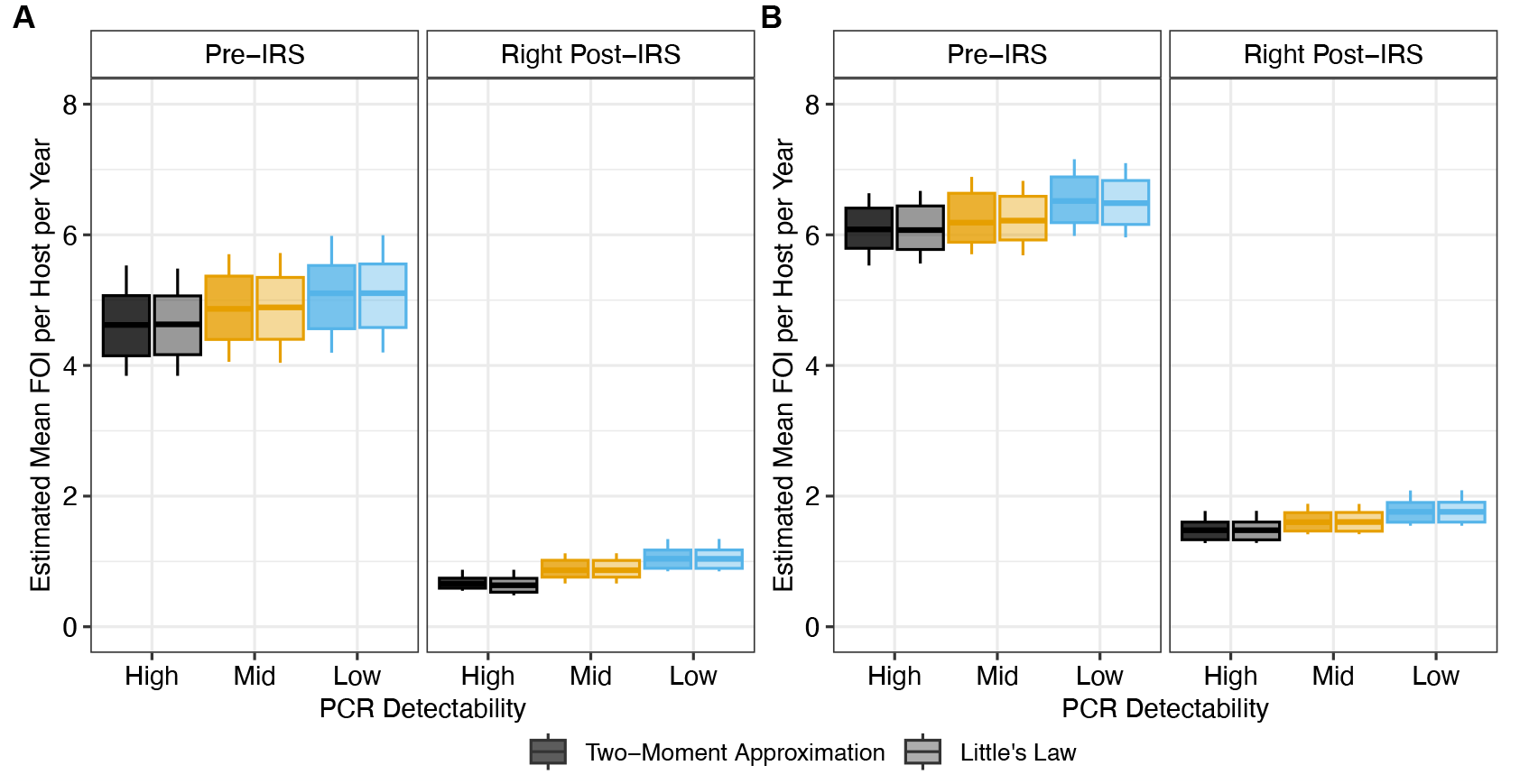
Confidence intervals for the estimated mean FOI values in Ghana surveys before and immediately after a transient three-round IRS intervention (A) The estimated FOI values when excluding these treated individuals from the analysis. (B) The estimated FOI values when discarding the infection status and MOI estimates of treated individuals and sampling from non-treated ones. Each boxplot shows minimum, 5% quantile, median, 95% quantile, and maximum. The value of *c* is set to 60.

**Figure 26.**
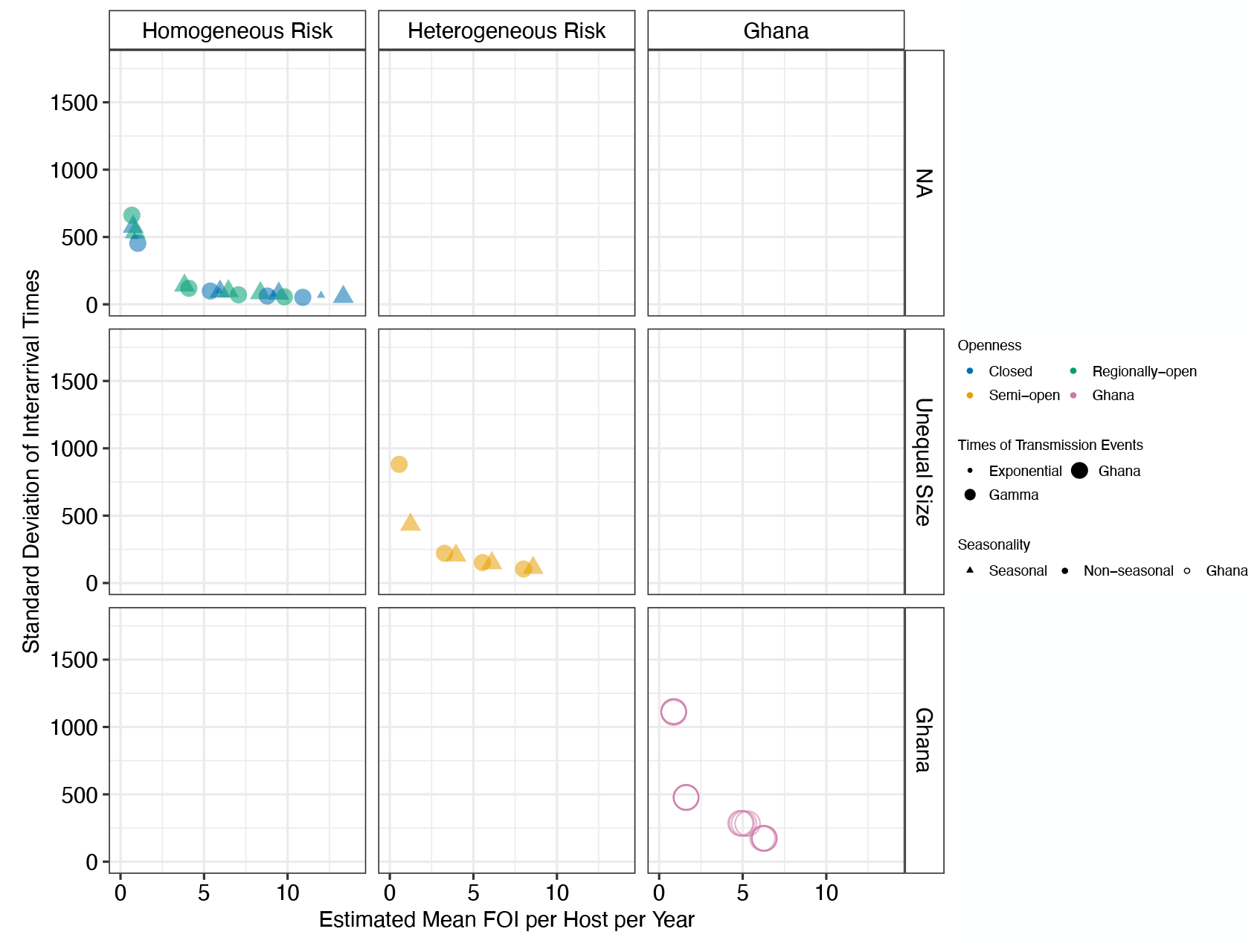
Estimated standard deviation of the inter-arrival times using the two-moment approximation method across different simulation scenarios and field data from Bongo District, Ghana.

**Figure 27.**
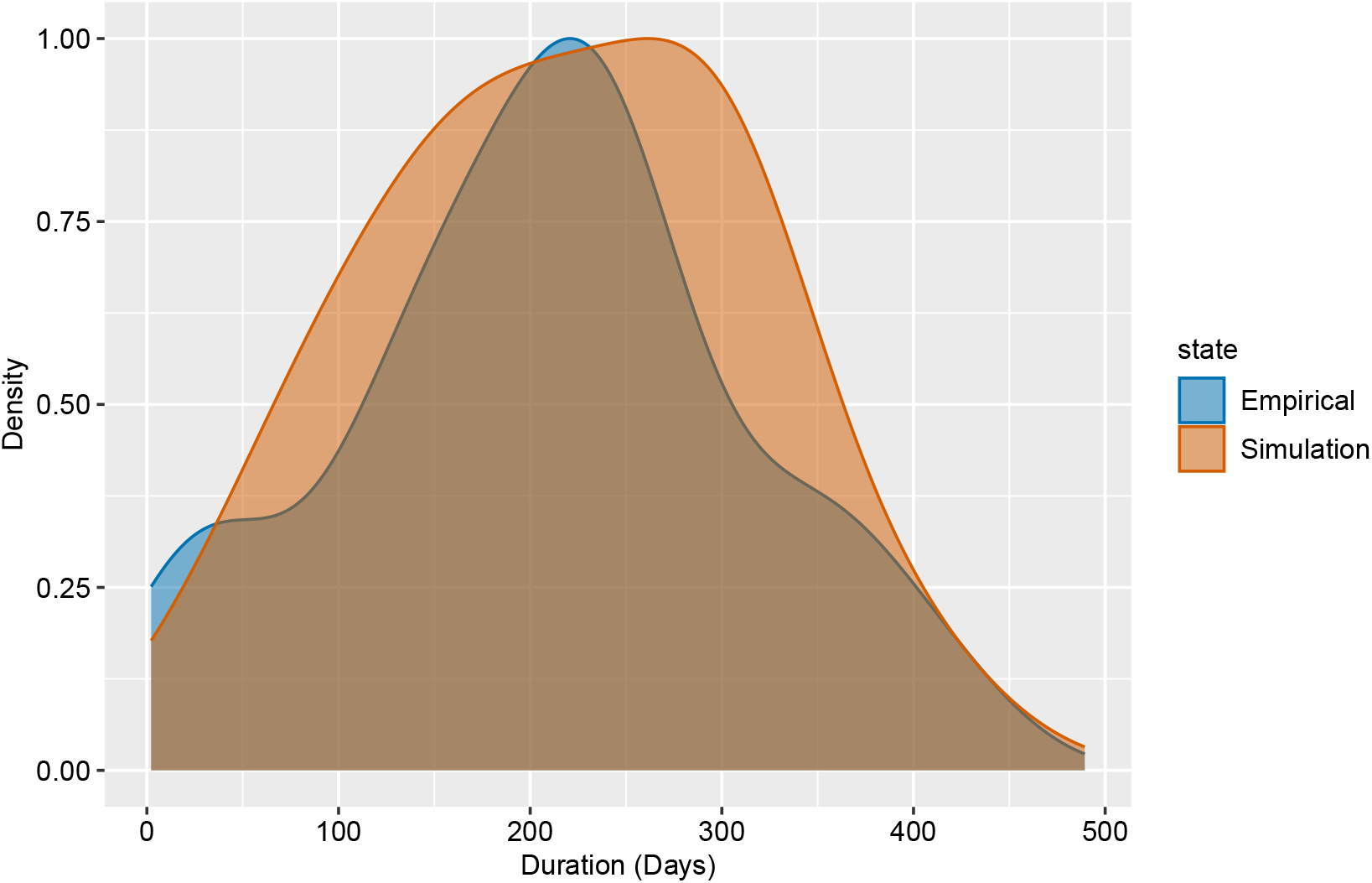
Comparison of the distribution of infection durations among naive hosts during the pre-IRS phase in simulated seasonal, semi-open systems where times between local transmission events follow a Gamma distribution, versus historical clinical data from neurosyphilis patients treated with *Plasmodium falciparum*. In the simulations, each infection can clear before all of its *var* genes have been expressed and recognized. Specifically, during the expression of any gene, whether non-final or final, there is a small probability of infection clearance that depends on the host’s pre-existing immunity to that gene’s epitopes (Appendix 1–Simulation data, subsection “An extended *var* model,” sub-subsection “Within-host dynamics”).

**Figure 28.**
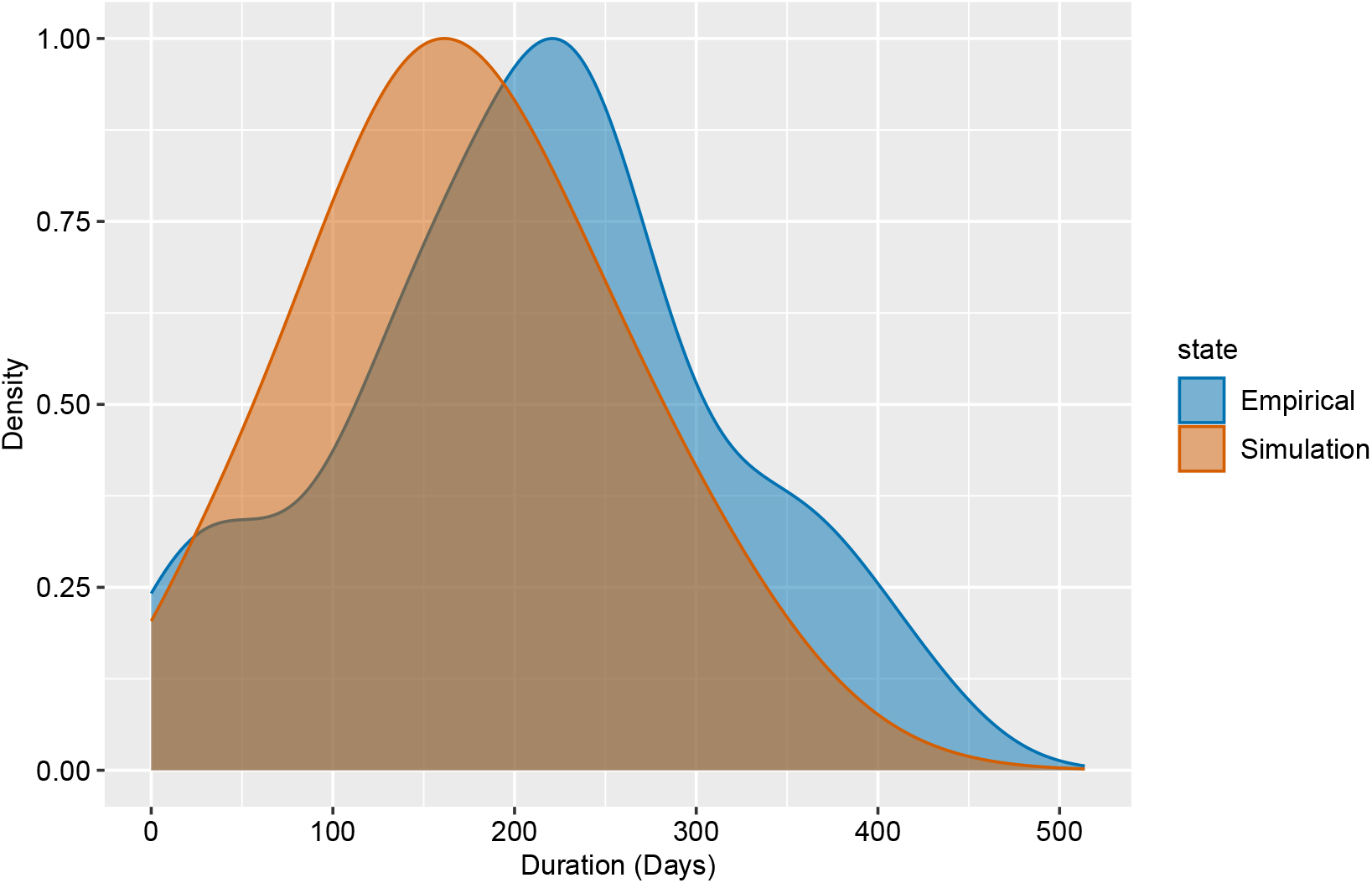
As in Appendix 1-Figure 27, we compare here the distribution of infection durations for the same simulation conditions with those from the historical clinical data, but show the results for children aged 1-5 years rather than naive hosts in the simulation.

**Figure 29.**
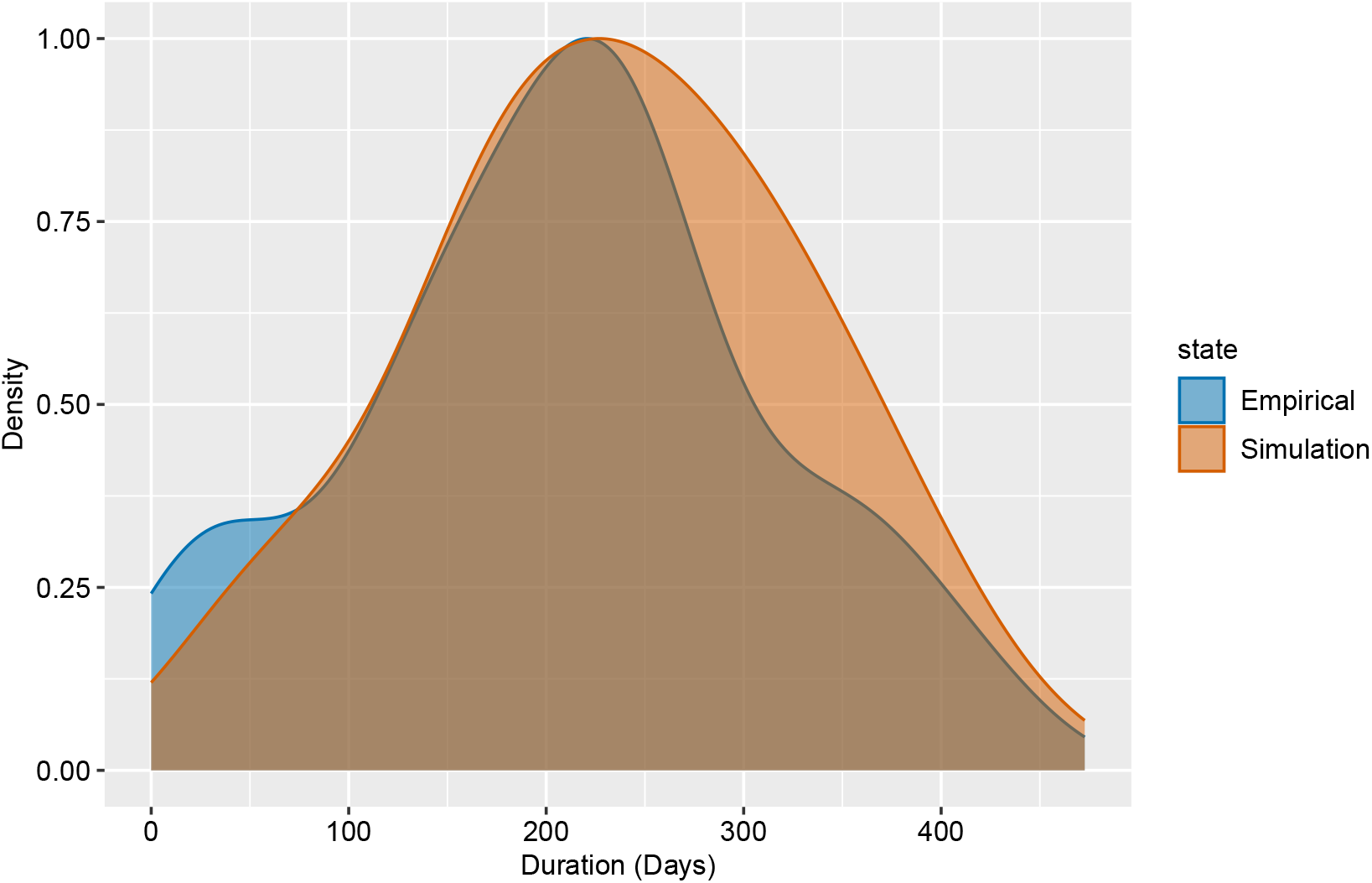
Comparison of the distribution of infection durations among naive hosts during the pre-IRS phase in simulated non-seasonal, semi-open systems where times between local transmission events follow a Gamma distribution, versus historical clinical data from neurosyphilis patients treated with *Plasmodium falciparum*. In the simulations, each infection can clear before all of its *var* genes have been expressed and recognized. Specifically, during the expression of any gene, whether non-final or final, there is a small probability of infection clearance that depends on the host’s pre-existing immunity to that gene’s epitopes.

**Figure 30.**
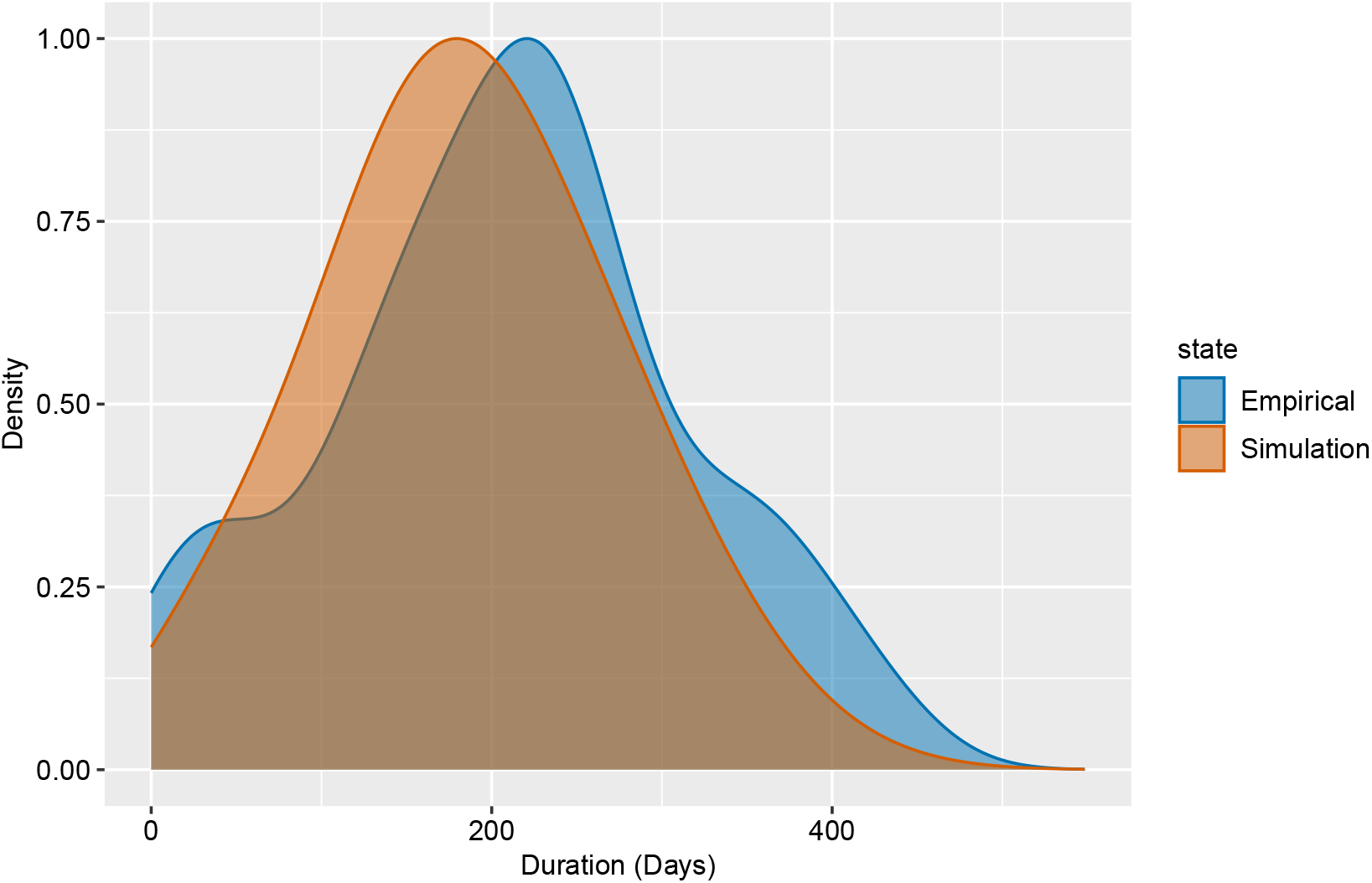
As in Appendix 1-Figure 29, we compare here the distribution of infection durations under the same simulation conditions with those from the historical clinical data, but show the results for children aged 1-5 years rather than naive hosts in the simulation.

**Figure 31.**
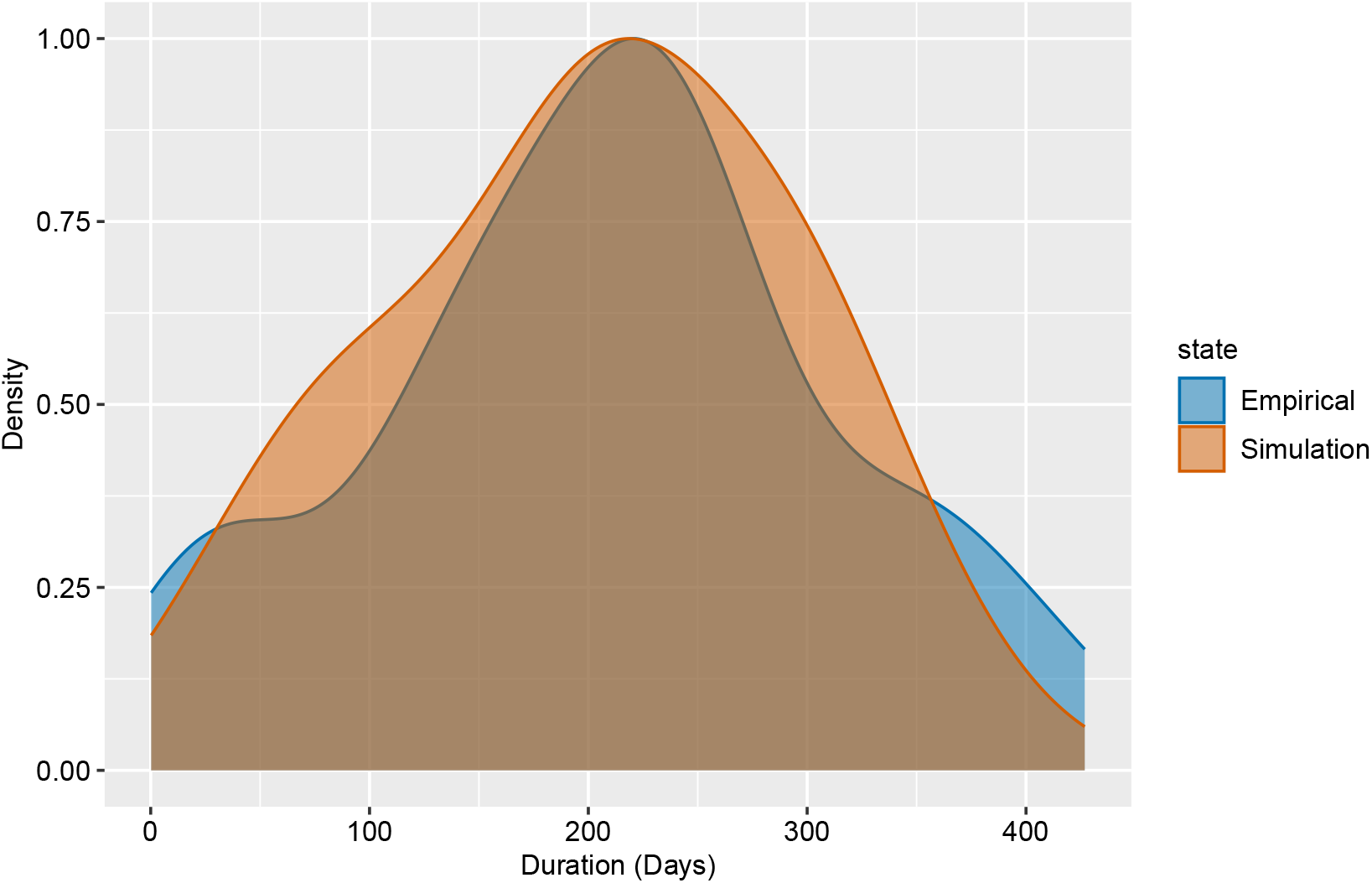
Comparison of the distribution of infection durations among naive hosts during the pre-IRS phase in simulated non-seasonal, regionally-open systems where times between local transmission events follow a Gamma distribution, versus historical clinical data from neurosyphilis patients treated with *Plasmodium falciparum*. In the simulations, each infection can clear before all of its *var* genes have been expressed and recognized. Specifically, during the expression of any gene, whether non-final or final, there is a small probability of infection clearance that depends on the host’s pre-existing immunity to that gene’s epitopes.

**Figure 32.**
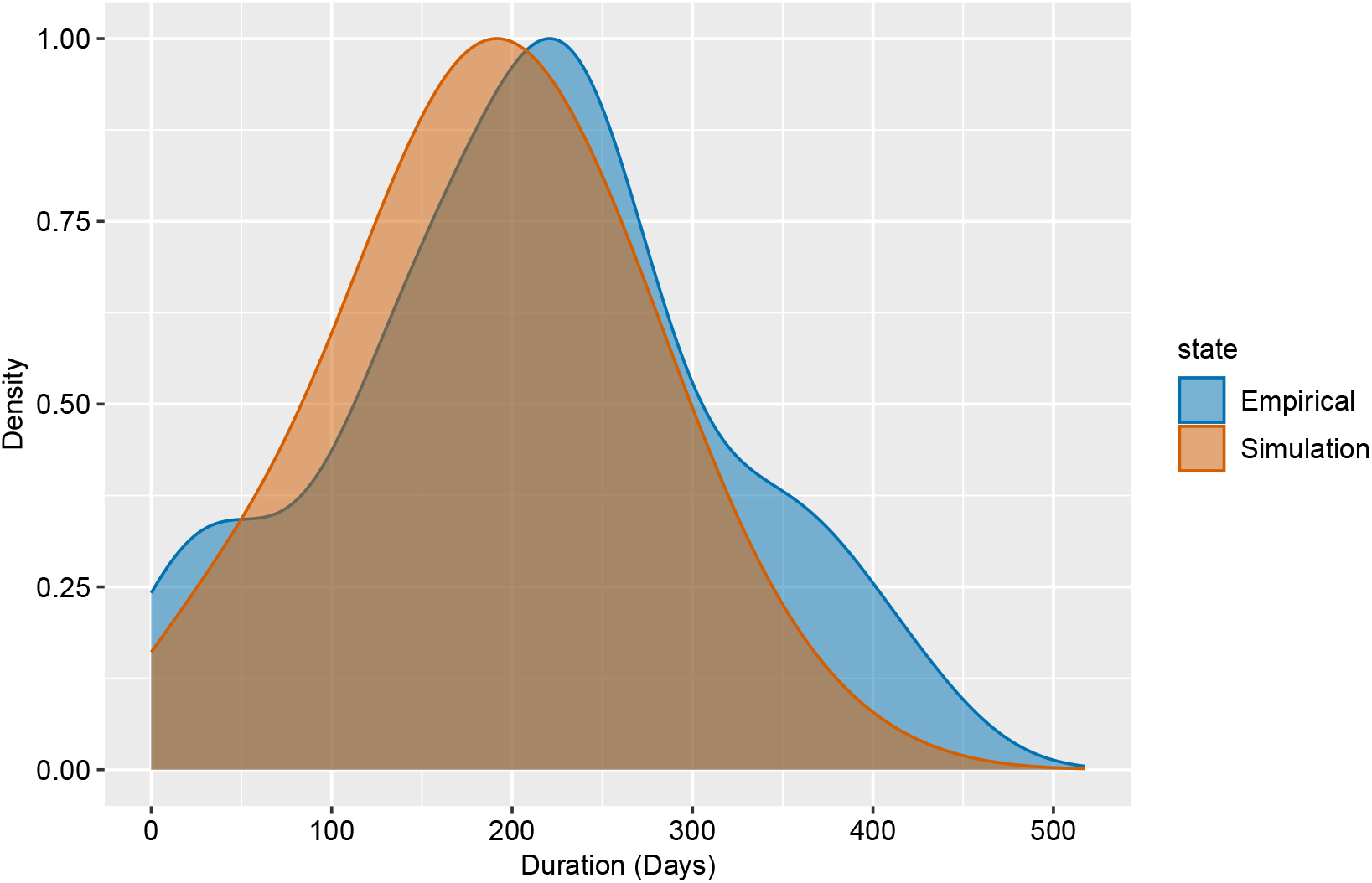
As in Appendix 1-Figure 31, we compare here the distribution of infection durations under the same simulation conditions with those from the historical clinical data, but show the results for children aged 1-5 years rather than naive hosts in the simulation.

## Data and code availability

The sequences utilized in this study are publicly available in GenBank under BioProject Number: PRJNA 396962. All data associated with this study, including de-identified individual participant data, are available in the manuscript, appendices, and on GitHub (https://github.com/UniMelb-Day-Lab/FOI_Pf_Ghana). Redistribution or reuse of these data requires proper attribution and prior approval. Researchers interested in further use of these data should contact the Malaria Reservoir Study Team, represented by the corresponding author, Prof. Karen Day, to discuss how these data will be utilized for academic or research purposes and, if appropriate, to identify opportunities for collaboration. The simulation code and analysis scripts are available at https://github.com/qzhan321/FOI.

## Acknowledgments

We wish to thank the participants, communities, and the Ghana Health Service in Bongo District, Ghana, for their willingness to participate in the study of empirical data. We would like to thank the field teams in Bongo for their technical assistance in the field, as well as the laboratory personnel at the Navrongo Health Research Centre for their expertise and for undertaking the sample collections and parasitological assessments. This research was supported by Fogarty International Center at the National Institutes of Health through the joint NIH-NSF-NIFA Ecology and Evolution of Infectious Diseases award R01-TW009670 to K.P.D. and M.P.; and the National Institute of Allergy and Infectious Diseases, National Institutes of Health through the joint NIH-NSF-NIFA Ecology and Evolution of Infectious Diseases award R01-AI149779 to K.P.D. and M.P. We appreciate the support of the Research Computing Center at the University of Chicago through the computational resources of the Midway cluster.

